# Considering social risk alongside genetic risk for bipolar disorder in the All of Us Research Program

**DOI:** 10.64898/2026.04.06.26349528

**Authors:** Rachel R. Sharp, Micah Hysong, Robert G. Mealer, Laura M. Raffield, LaShaunta Glover, Michael I. Love

## Abstract

Polygenic risk scores (PRS) have shown increasing utility for risk stratification across complex diseases, but for psychiatric disorders such as bipolar disorder (BD), current PRS explain only a fraction of disorder liability (∼1-9%), with predictive performance further diminished in non-European populations and real-world clinical cohorts. To explore the potential of integrating social and environmental risk factors alongside genetic liability to improve risk prediction, we evaluated the relationship between a PRS for BD (PRS_BD_) and six social risk measures – perceived stress, discrimination in medical settings, neighborhood social cohesion, perceived neighborhood disorder, cost-related medication nonadherence, and adverse childhood experiences – to BD case status in 115,275 participants (7,000 cases; 108,275 controls) from the All of Us Research Program. PRS_BD_ was associated with BD case status across ancestry groups, though liability-scale variance explained was attenuated relative to what has been reported for curated research cohorts (R² = 1.86% in European, 0.60% in African, 1.65% in Latino/Admixed American ancestries). Each social risk factor tested exhibited a larger effect size than PRS_BD_, with perceived stress (OR = 2.05 per SD) and adverse childhood experiences (OR = 2.68 for ≥4 ACEs) demonstrating the strongest associations. Individuals in the lowest genetic risk decile with high social burden exhibited BD prevalence comparable to or exceeding those in the highest genetic risk decile with low social burden. These findings demonstrate the substantial explanatory power of social risk factors and support the development of integrated genetic-social risk frameworks for more accurate and equitable psychiatric risk prediction.

## Introduction

Neuropsychiatric conditions, including major depression, anxiety, and bipolar disorder (BD), affect an estimated 1 in 5 adults in the U.S. each year and are leading contributors to the global burden of disease.^1,2^ In recent decades, the psychiatric genetics community has made substantial progress identifying the genetic contributions of these disorders.^3–6^ Yet for many of these conditions, common genetic factors explain only a fraction of disorder risk, motivating efforts to improve risk prediction by integrating genetic information with environmental and social context.^7–9^

Polygenic risk scores (PRS) aggregate estimated effects of common trait-associated variants across the genome into a single value, representing the additive component of an individual’s genetic liability for the trait. Although PRS have shown increasing utility in risk stratification across a number of diseases,^10,11^ current PRS capture only a fraction of total heritability for psychiatric disorders. For BD, SNP-based heritability estimates reach ∼22%, well below twin-based estimates of ∼60-80%, and PRS explain only ∼9% of phenotypic variance on the liability scale.^12–16^ The gap between twin and SNP heritability may in part reflect contributions from genetic factors not captured by PRS, including rare coding and structural variants that may be particularly relevant for some psychiatric disorders.^17,18^ Moreover, PRS derived from predominantly European-ancestry discovery samples show substantially reduced predictive performance in non-European-ancestry populations due to differences in linkage disequilibrium structure and allele frequencies, a limitation that remains a major barrier to equitable clinical implementation.^19^

The likelihood of developing a polygenic disorder such as BD is not solely influenced by genetic factors but also by social, environmental, and economic risk factors, collectively referred to as social determinants of health (SDOH).^20^ Several such factors have been associated with BD onset and severity. Childhood maltreatment or adversity has been consistently linked to earlier onset and disorder severity, though whether this relationship is moderated by polygenic burden remains unclear, with some studies reporting gene-environment interactions,^21–23^ and others finding primarily additive effects.^24^ Stressful life events have been associated with mood episode onset in genetically at-risk individuals, and socioeconomic deprivation has been robustly associated with the development and persistence of mood disorders.^25,26^ Additionally, experiences of discrimination in medical settings may contribute to diagnostic disparities, with evidence suggesting that Black Americans with BD are disproportionately misdiagnosed with schizophrenia, potentially reflecting both clinician bias and differential access to psychiatric care.^27,28^

Aside from specific associations with BD, promising results indicate improved risk prediction and phenotypic discrimination when modifiable risk factors are included in models alongside genetic risk scores,^29–31^ and researchers have increasingly called for the inclusion of SDOH in risk prediction models across traits.^32–34^ Methodologically, omission of strong social covariates from these models can bias risk estimates regardless of whether the omitted factors correlate with the PRS or modify its association strength.^35^ Yet how the effects of genetic and social risk factors combine, or potentially interact, to contribute to complex disorder risk remains poorly understood.^36,37^ Particularly in neuropsychiatry, rigorous evaluation of how social and environmental factors modulate disorder susceptibility is required before PRS can be adopted for clinical use.^7,38,39^

Here we evaluate the contributions of social risk and genetic risk to the development of BD in a diverse U.S. population. We leverage data from the *All of Us Research Program* (AoU) to assess the predictive power of an externally developed PRS for BD (PRS_BD_) derived from the latest PGC meta-analysis, alongside several social risk measures derived from AoU surveys. We model the effects of genetic and social risk factors and test for potential interactions to quantify their individual and combined contributions to BD case status. By integrating genetic liability with social context, we seek to improve risk prediction strategies for complex psychiatric traits and underscore the value of incorporating social determinants into genetic risk frameworks for a holistic approach to psychiatric epidemiology.

## Subjects and methods

### All of Us Cohort

We utilized data from the *All of Us* (AoU) Research Program, a diverse cohort aiming to recruit one million participants across the United States, a detailed report of which, including objectives, data availability, and limitations, has been previously published.^40^ This study utilized data from 414,830 AoU participants with short-read whole-genome sequencing (srWGS) data enrolled by October 1, 2023 (Data Release Version v8) and was conducted under the AoU Data Use Agreement.

Of the 414,830 participants with linked srWGS data, 115,275 participants were ultimately included in statistical analyses following genomic QC, EHR filtering, and application of our phenotyping schema (Figure 1). Included participants had linked demographic and Electronic Health Record (EHR) data and had completed the relevant AoU survey modules for the investigated social risk variables (at missingness thresholds defined below). To ensure sufficient health record density for subsequent analyses, we restricted the cohort to participants with an EHR record length of 3 years or more and a record depth of 3 or more distinct clinical visits. Finally, we restricted the final cohort to individuals with predicted genetic ancestry of either European, African, Admixed American, or East Asian descent (matching the GWAS source ancestries), who were aged 20 to 69 years at their last recorded event. South Asian and Middle Eastern predicted ancestry groups were excluded from all analyses due to case counts below 20 in each group.

**Figure 1.**
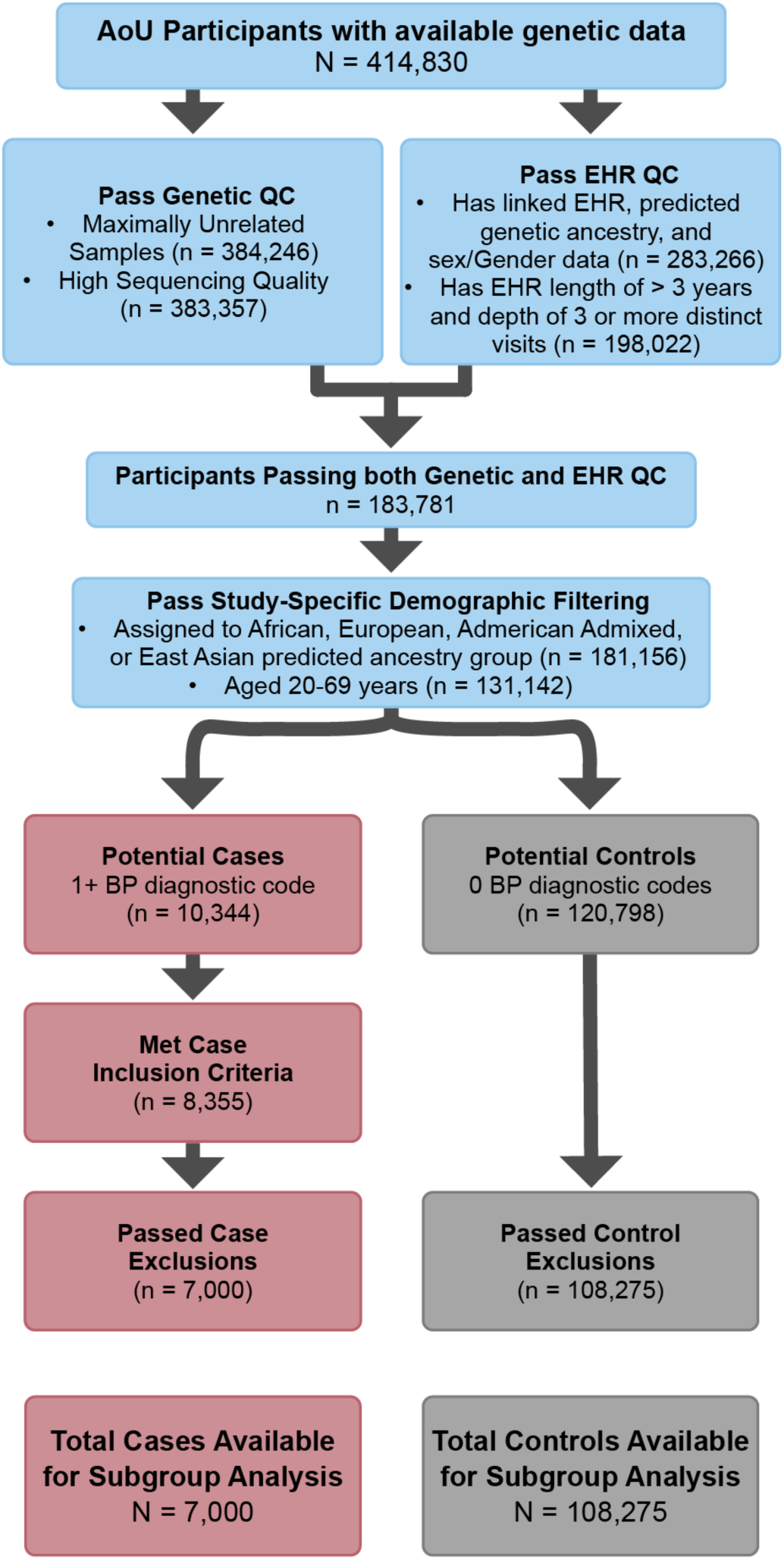
Flowchart detailing derivation of analysis cohort. Flowchart illustrating sample quality control, participant inclusion criteria, and EHR phenotyping steps taken to produce the final analysis cohort.

### Whole-Genome Sequencing Data

We utilized the centrally processed v8 genotype data provided by the AoU Researcher Workbench. Specifically, we used the ACAF-thresholded PLINK-formatted files derived from short-read whole genome sequencing data, which retain variants exceeding a population-specific allele count of 100, or a population-specific allele frequency of 1%.

We removed samples flagged by AoU as having low-quality WGS data during the joint callset QC process. To account for cryptic relatedness, we utilized the pre-computed kinship scores provided by AoU and excluded the AoU-recommended list of samples to generate a maximally unrelated set of participants.^41^

### Demographic and Ancestry Variables

We defined a three-level sex/gender variable based on concordance between sex assigned at birth and self-reported gender identity. Participants were classified as Cis-Male (Assigned male at birth, identified gender as male), Cis-Female (assigned female at birth, identified gender as female), or Sexual and Gender Minority (SGM) (discordant sex and gender indications, intersex, or non-binary identities). Participant age was calculated as the interval between the date of birth and the date of the last recorded event in their EHR data. EHR length and record depth were derived from available EHR data, calculating EHR length as the interval between the first and last recorded event and record depth as the number of distinct recorded events. Finally, genetic ancestry, as well as genetic ancestral Principal Components (PCs), was defined using the pre-computed ancestry predictions provided by AoU as previously described.^41^

### PRS Calculation

We computed polygenic risk scores for BD (PRS_BD_) in our cohort using publicly available summary statistics from the largest multi-ancestry Genome-wide Association Study (GWAS) of BD to date, performed by the Psychiatric Genomics Consortium.^12,42^ Specifically, we utilized summary statistics from the meta-analysis multi-ancestry analysis excluding self-reported cases (N_cases_ = 67,948; N_controls_ = 867,710). We selected the multi-ancestry GWAS without self-report as O’Connell et al. demonstrated that self-reported cases have a distinct genetic architecture with substantially lower SNP-heritability (h^2^_SNP_ = 0.08) compared to clinically ascertained cases (h^2^_SNP_ = 0.22), and that PRS derived from this sub-cohort showed better predictive performance in clinical cohorts (Weighted Liability R^2^ = 0.090) compared to models including self-reported data (R^2^ = 0.058).

Following the approach used in O’Connell et al., we computed PRS for our cohort using the Bayesian regression framework PRS-CS-auto to estimate posterior SNP effect sizes.^43^ While the source GWAS was a multi-ancestry meta-analysis, a majority of participants (∼82%), were of European descent. Consequently, we elected to use the 1000 Genomes Project Phase 3 European Reference panel provided by the PRS-CS developers to ensure sample independence.^44^ PRS-CS-auto was run on each chromosome with 10,000 MCMC iterations and 5,000 burn-in steps, as recommended by developers.^43^

Posterior effect sizes were lifted over from GRCh37 to GRCh38 using Hail (v0.2.130)^45^. We performed allele harmonization by removing strand-ambiguous variants and aligning effect alleles to the target dataset, flipping the sign of the effect size where the effect allele matched the reference. Variants failing liftover or with unresolved allele mismatches were excluded. Individual-level polygenic risk scores were calculated using PLINK v1.9^46,47^ with the --score function, summing the dosage of the effect allele weighted by the posterior effect size estimated by PRS-CS-auto. PRS-CS-auto estimated posterior effect sizes for 1,061,483 variants. After liftover to GRCh38 (414 variants failed), removal of strand-ambiguous variants, and allele harmonization with the AoU target dataset, 1,045,187 variants were retained for score calculation.

To address potential effects of population stratification, raw PRS values were adjusted for the top 16 genetic ancestry PCs.^48^ The resulting residuals were standardized (mean=0, SD=1) within the full cohort to generate the final global PRS used for analysis. We confirmed that this residualization of the PRS score on a continuous measure of ancestry largely removed large population-scale differences in the PRS distributions (Figure S1).

### Bipolar Disorder Phenotyping

Bipolar disorder (BD) case and control status was determined using EHR data within the AoU Researcher Workbench. Cases were defined as participants meeting one of the following criteria: (1) presence of at least 2 diagnostic codes (ICD-9/10 or SNOMED) for BD more than 30 days apart; (2) presence of at least 1 BD diagnostic code and a prescription record for a BD-indicated medication within one year of each other; or (3) presence of at least 1 BD diagnostic code and a high volume of BD-indicated medications (≥30 total records or ≥3 unique medications). BD-indicated medications included Lithium, Valproate, Lamotrigine, and Carbamazepine (for a detailed list, see Supplemental Information). For participants meeting at least one of the above criteria, we next excluded those with codes for a conflicting psychiatric condition (e.g., schizophrenia) that outnumbered the participant’s BD code count, unless the BD codes were most recent. This case definition strategy was adapted from a recent clinician-informed EHR-based phenotyping approach for early-onset bipolar disorder.^49^ Requiring convergent evidence from repeated diagnostic codes or concordant medication records provides a more stringent case definition than single-code approaches, which have been shown to capture heritable variation unrelated to the target disorder.^50^ Controls were defined as participants with no BD diagnostic codes and no exclusionary psychiatric diagnoses. For full code and medication lists used to construct the cohort, see Supplemental Information.

To assess the consistency of our EHR-based case definitions, we examined the concordance with self-reported personal and family history of BD. Specifically, we calculated PRS risk curves across subgroups of participants who self-reported (1) having at least one parent with BD, (2) having BD themselves, (3) currently receiving medication or treatment for BD, and (4) currently seeing a doctor for BD. We additionally compared rates of self-reported BD history between EHR-defined cases and controls. While self-report measures are subject to recall bias and respondents may be unable to reliably discriminate between psychiatric diagnoses,^51^ the concordance observed between EHR-based and self-report classifications provides additional support for the reliability of our phenotyping strategy.

### Social Determinants of Health Measures

We selected six social risk measures spanning domains with prior evidence of association with BD or related psychiatric outcomes. Perceived stress has been associated with mood episode onset in genetically at-risk individuals,^52,53^ experiences of discrimination in medical settings may contribute to diagnostic disparities and case misclassification in BD,^27^ cost-related medication non-adherence captures economic barriers to treatment continuity,^54,55^ a factor linked to poor psychiatric outcomes, adverse childhood experiences have been consistently associated with earlier BD onset and greater illness severity,^22,56,57^ and neighborhood social cohesion and perceived neighborhood disorder capture community-level social and physical environment exposures associated with psychiatric morbidity.^58–60^

Measures were derived from participant-provided responses to “The Basics,” “Social Factors of Health,” “Health Care Access and Utilization Survey,” “Personal/Family Health History,” and “Emotional Health History and Well-being” surveys. From these, individual survey responses were extracted and scored according to validated guidelines for each instrument. These six surveys were: Perceived Stress (PSS-10),^61,62^ Discrimination in Medical Settings (DMS),^63^ adapted from the Everyday Discrimination Scale,^64^ Cost-Related Medication Non-adherence (CRMN),^65^ Neighborhood Social Cohesion (SCNS),^66^ Perceived Neighborhood Disorder (PNDS),^67^ and Adverse Childhood Experiences (ACE).^68^

The following analyses were performed separately for each survey. First, participants who completed less than 70% of the items for a specific survey were excluded from the analysis for that survey. Patterns of item-level missingness were visualized and assessed using the *naniar* R package (Figure S2A).^69^ For the remaining participants, missing item-level values were imputed using a k-nearest neighbors approach (k=5) implemented using the *caret* R package prior to scoring.^70^ Finally, distributional accuracy of score imputation was assessed by comparing imputed distributions to complete-case distributions (Figure S2B).

Prior to association analysis, survey scores were transformed for comparison. Inter-scale correlations among the six social risk measures are shown in Figure S2C. Continuous scores (PSS, DMS, SCNS, and PNDS) were standardized to Z-scores (Mean=0, SD=1). Directionality was standardized such that higher scores reflect higher risk. CRMN and ACE scores were analyzed as binary variables with a CRMN value of 1 indicating any behavior of Cost-Related Medication Nonadherence, and an ACE value of 1 indicating 4 or more adverse experiences.^68^

Due to varying completion rates across surveys and ancestry groups, analytic sample sizes differed for each social risk variable (range: N=17,098-64,009). Full sample sizes by survey and ancestry group are reported in Table S1, and completion rates by case/control status and ancestry are shown in Figure S3.

### Statistical Analyses

All statistical analyses were performed on the AoU Researcher Workbench using R version 4.4.0.^71^ Data manipulation was conducted using tidyverse, and descriptive statistics and cohort characteristics tables were generated using the gtsummary and flextable packages.^72–74^ We assessed the performance of the PRS using logistic regression models adjusted for age (linear and quadratic terms), sex/gender, EHR record length and depth, and 16 genetic PCs. Model discrimination was evaluated using the Area Under the Curve (AUC) calculated using the pROC package.^75^ Variance explained was estimated using liability-scale R^2^ assuming a population prevalence of 2%^12^ with 95% CIs estimated via bootstrapping (1,000 replicates) implemented using the boot package.^76,77^

For each social risk variable, we used logistic regression to model BD case status as a function of polygenic risk score, the social risk variable, age, age^2^, sex/gender, the first 16 genotype PCs, EHR record length, and record depth. All PRS associations were estimated using the continuous, standardized PRS. Decile-based analyses are presented for visualization purposes only. Model outputs were processed using the broom package.^78^ To assess potential multiplicative effects, we fitted models including a PRS X Social Risk interaction term. We tested for multiplicative interaction using a likelihood ratio test (LRT) comparing the interaction model to a base model without the interaction term. Statistical significance was assessed after correcting for multiple testing using the Benjamini-Hochberg procedure. All PRS performance, social risk association, and interaction models were fit in the combined “All Ancestries” sample and separately within European, African, and Admixed American ancestry groups. East Asian participants were included in the All Ancestries analysis but excluded from stratified models due to insufficient sample size (see demographic and ancestry variables).

## Results

### Study design and cohort characteristics

Out of 414,830 participants in the AoU v8 cohort with available srWGS data, a total of 115,275 individuals (7,000 cases and 108,275 controls) were retained for further analyses after applying QC and cohort selection criteria (Figure 1). The cohort was predominantly cis-female (66%), with a mean age of 50.5 years (SD = 13.2) (Table 1). From here, we stratified further analyses by genetically predicted ancestry group as defined by the centrally processed AoU ancestry predictions. As expected, most participants were of European ancestry (4,054 cases and 63,370 controls), followed by participants of African (1,953 cases and 24,014 controls) and Latino/Admixed American ancestry (944 cases and 19,271 controls). East Asian participants were ultimately excluded from ancestry-stratified analyses due to low sample size (49 cases and 2,618 controls) but were retained in the “All ancestries” data. Notably, most subsequent analyses utilize smaller sub-samples due to varying completion of the necessary social risk surveys; analytic sample sizes ranged from N=17,098 (ACE) to N=64,009 (CRMN), with four of six surveys yielding samples of approximately 49,000. Full sample sizes by survey and ancestry group are reported in Table S1. Because participants were included in a given survey’s analysis only if they completed at least 70% of that survey’s items, analytical sub-sample sizes varied across surveys. The proportion of participants meeting this threshold differed by survey, ancestry group, and case status (Figure S3), with European ancestry participants generally showing higher completion rates and a notably larger difference in completion rate between cases and controls than was observed in African and Admixed American ancestry groups.

**Table 1.**
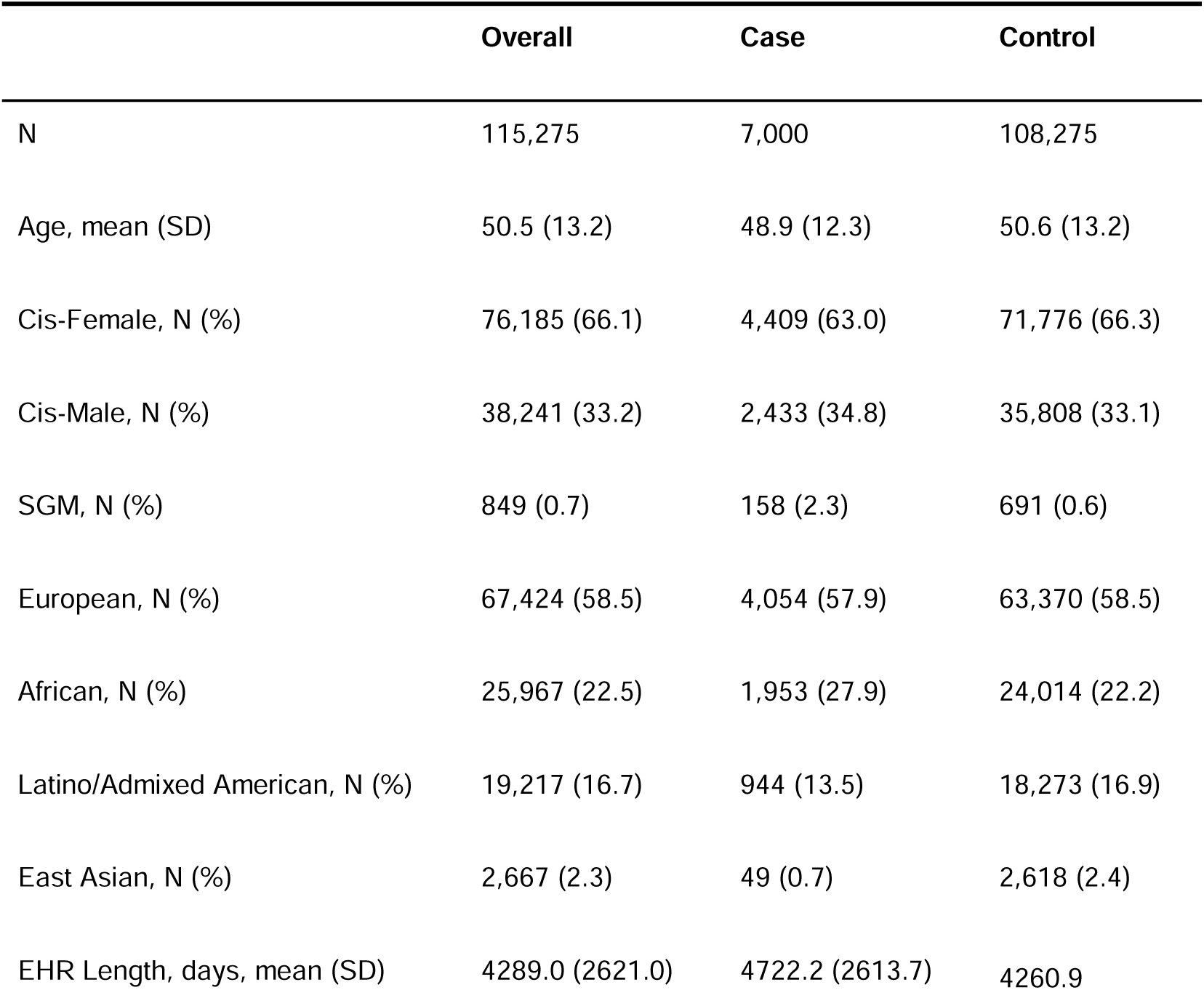
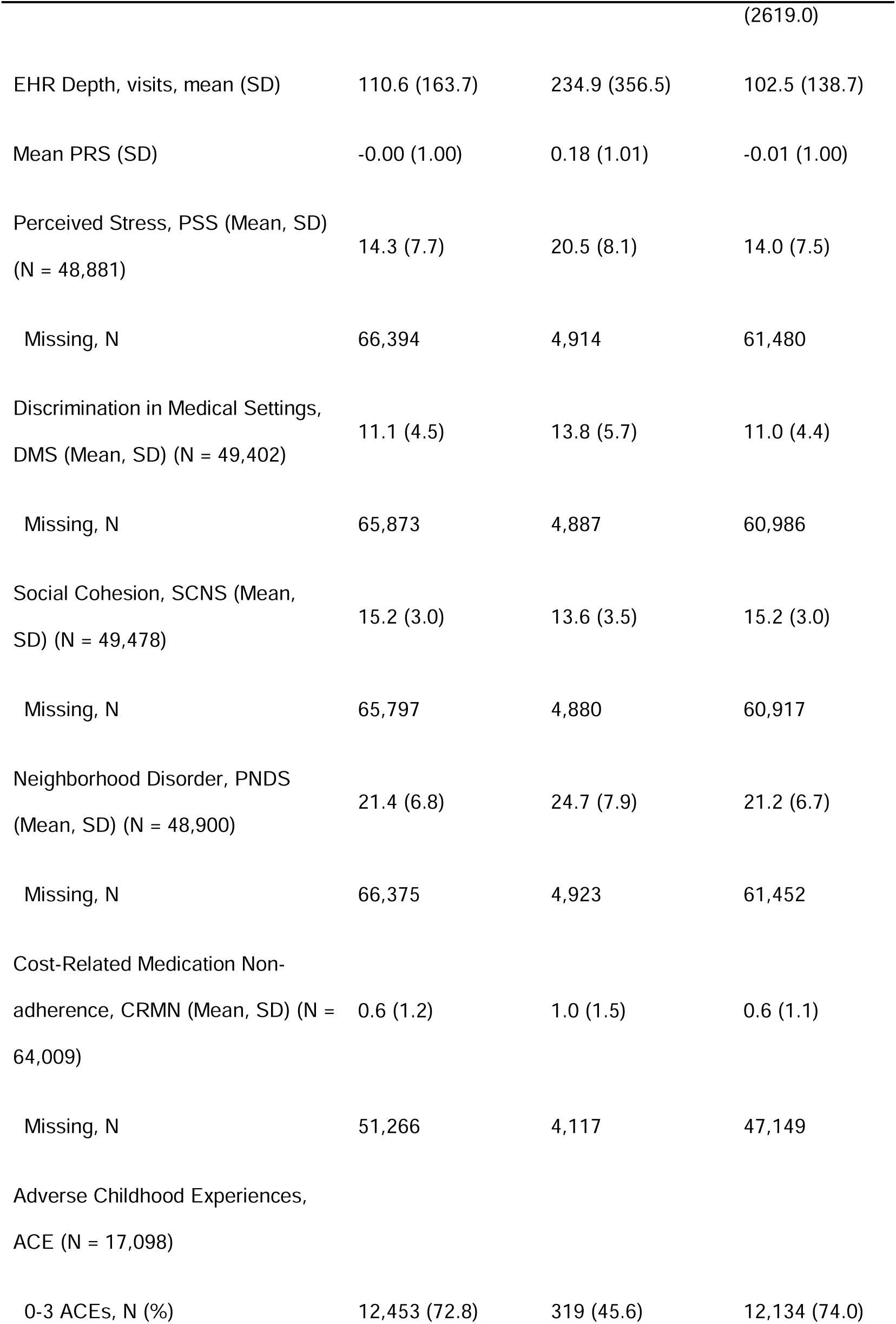
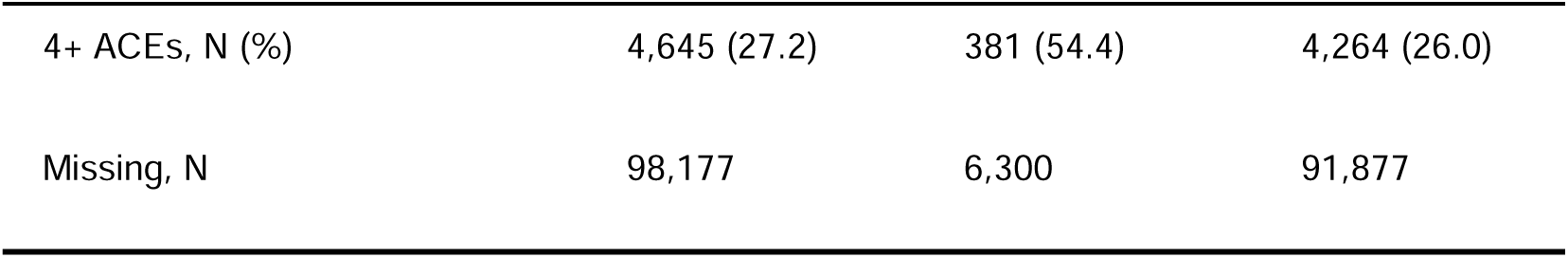
Cohort Demographic and Social Risk Characteristics.

### Polygenic Risk Score Performance

We computed individual polygenic risk scores (PRS_BD_) in our sample of 7,000 BD cases and 108,275 controls, observing a positive association between PRS_BD_ and odds of BD case status across ancestries, though predictive performance varied by population (Table 2; Figure 2). In the All Ancestries analysis, each standard deviation increase in PRS_BD_ was associated with a 22% increase in odds of BD case status (OR = 1.22; 95% CI 1.19-1.25; P = 5.91 × 10^-52^). Liability-scale R^2^ was highest in the European group (1.86%; 95% CI 1.29–2.47), followed by Latino/Admixed American (1.65%; 95% CI 0.68–3.16), with lower performance in the African ancestry group (0.60%; 95% CI 0.22–1.12). These values are substantially attenuated relative to the PGC target cohorts, where O’Connell et al. reported median R^2^_liability_ of 9.0% in European-ancestry groups 2.2% in African ancestry groups.^12^ In absolute terms, observed BD prevalence ranged from 4.4% in the lowest PRS decile to 8.6% in the highest decile in the All Ancestries sample (risk ratio = 2.0), with the strongest separation observed in the European subsample (4.2% vs. 8.8%; risk ratio = 2.1). Theoretical absolute risk curves illustrate the translational population-level risk estimates by ancestry, ranging from 1% to 5% across the PRS distribution (Figure S6).

**Figure 2.**
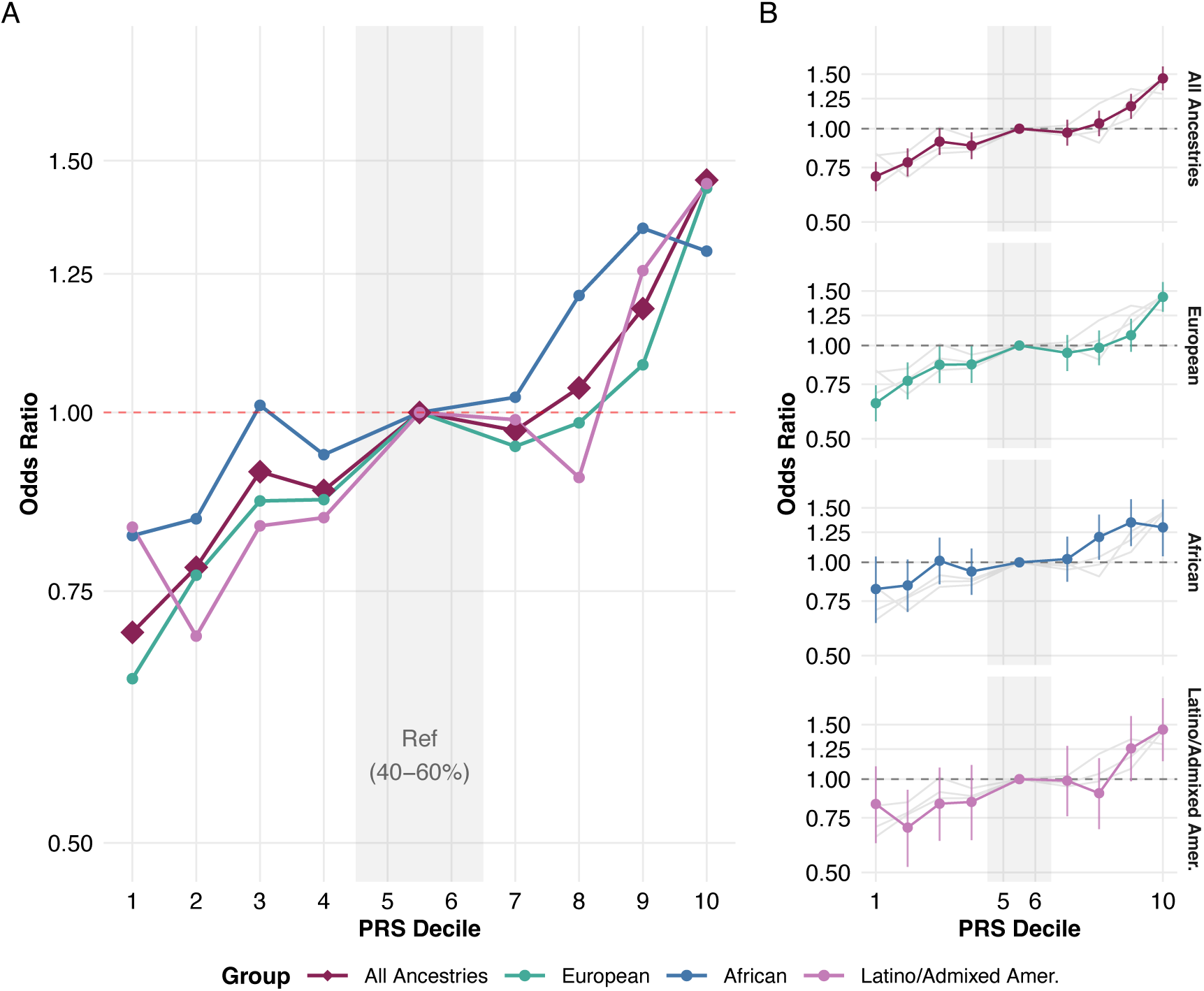
Bipolar disorder polygenic risk score risk stratification across ancestry groups. (A) Adjusted odds ratios (OR) of bipolar case status across PRS deciles. ORs were calculated relative to the middle quintile (deciles 5-6, indicated by the shaded region) using a logistic regression model adjusted for age, age^2^, sex/gender, EHR record length, record depth, and 16 genetic PCs. Trend lines are split by genetically predicted ancestry group, faceted out to the right. (B) Error bars represent the 95% confidence intervals (95% CI) derived from the adjusted logistic regression models. Grey background lines show the trends of other groups for comparison.

**Table 2.** Polygenic Risk Score Performance Across Ancestry Groups.

| Ancestry | N (Cases) | OR per SD (95% CI) | R<sup>2</sup> liability, % (95% CI) | P |
| --- | --- | --- | --- | --- |
| All Ancestries | 115,275 (7,000) | 1.22 (1.19–1.25) | 1.53 (1.15–2.01) | 5.91e-52 |
| European | 67,424 (4,054) | 1.22 (1.18–1.26) | 1.86 (1.29–2.47) | 1.00e-36 |
| African | 25,967 (1,953) | 1.18 (1.11–1.25) | 0.60 (0.22–1.12) | 1.31e-07 |
| Latino/Admixed American | 19,217 (944) | 1.21 (1.13–1.30) | 1.65 (0.68–3.16) | 4.56e-08 |

To evaluate model discrimination, we assessed AUC across three covariate sets: a minimally adjusted model matching O’Connell et al. (Set 1: sex + 5PCs), our primary fully adjusted model (Set 2: age + age^2^ + sex/gender + EHR record length + EHR depth + 16 genetic PCS), and models additionally including each social risk variables (Set 3). For each covariate set, we additionally computed change in AUC between the covariate-only model and the same model with PRS added, isolating the individual PRS contribution in each scenario. Across covariate sets, PRS contributed modestly to model discrimination, with change in AUC in Set 2 ranging from 0.002 to 0.009 across ancestry groups. Full results are reported in Table S3.

Despite the modest variance explained, we observed a consistent linear increase in odds of BD case status across PRS deciles (Figure 2). Relative to individuals at median risk (40^th^-60^th^ percentile), individuals in the top decile had elevated odds of BD in the combined analysis (OR = 1.45; 95% CI 1.33-1.59), with similar elevation seen in European (OR = 1.44; 95% CI 1.29-1.60), African (OR = 1.30; 95% CI 1.05-1.60), and Latino/Admixed American groups (OR = 1.45; 95% CI 1.14-1.83). Full decile-stratified odds ratios by ancestry are reported in Table S2.

We additionally examined concordance between our EHR-based phenotyping and self-reported personal and family history of BD (see methods). Across four self-report indicators, higher PRS was associated with higher probabilities of EHR-defined case status, and EHR-defined BD cases self-reported personal or family history of BD at substantially higher rates than controls (Figure S4). PRS distributions across EHR case assignment strategies are shown in Figure S5.

### Association of Social Risk Factors with Bipolar Disorder

We next evaluated the relationship between six social risk factors and BD case status. We note that survey completion rates differed across cases and controls (Figure S3), thus the social Odds Ratios (ORs) reported here represent associations specific for this subsampled cohort and cannot easily be generalized to the larger population.

In our sample, however, we observed significant associations between higher social risk and odds of BD case status across all ancestry groups (Figure 3). Among the continuous variables tested, PSS yielded the largest effect size; in the All Ancestries analysis, every one SD increase in PSS was associated with an approximately 2-fold increase in the odds of BD case status (OR = 2.05; 95% CI 1.96-2.15). Similarly, for every one SD increase in SCNS, we observed an approximately 1.5-fold increase in the odds of BD case status (OR = 1.55; 95% CI 1.49-1.62), with DMS (OR = 1.47) and PNDS (OR = 1.49) showing comparable effect sizes.

**Figure 3.**
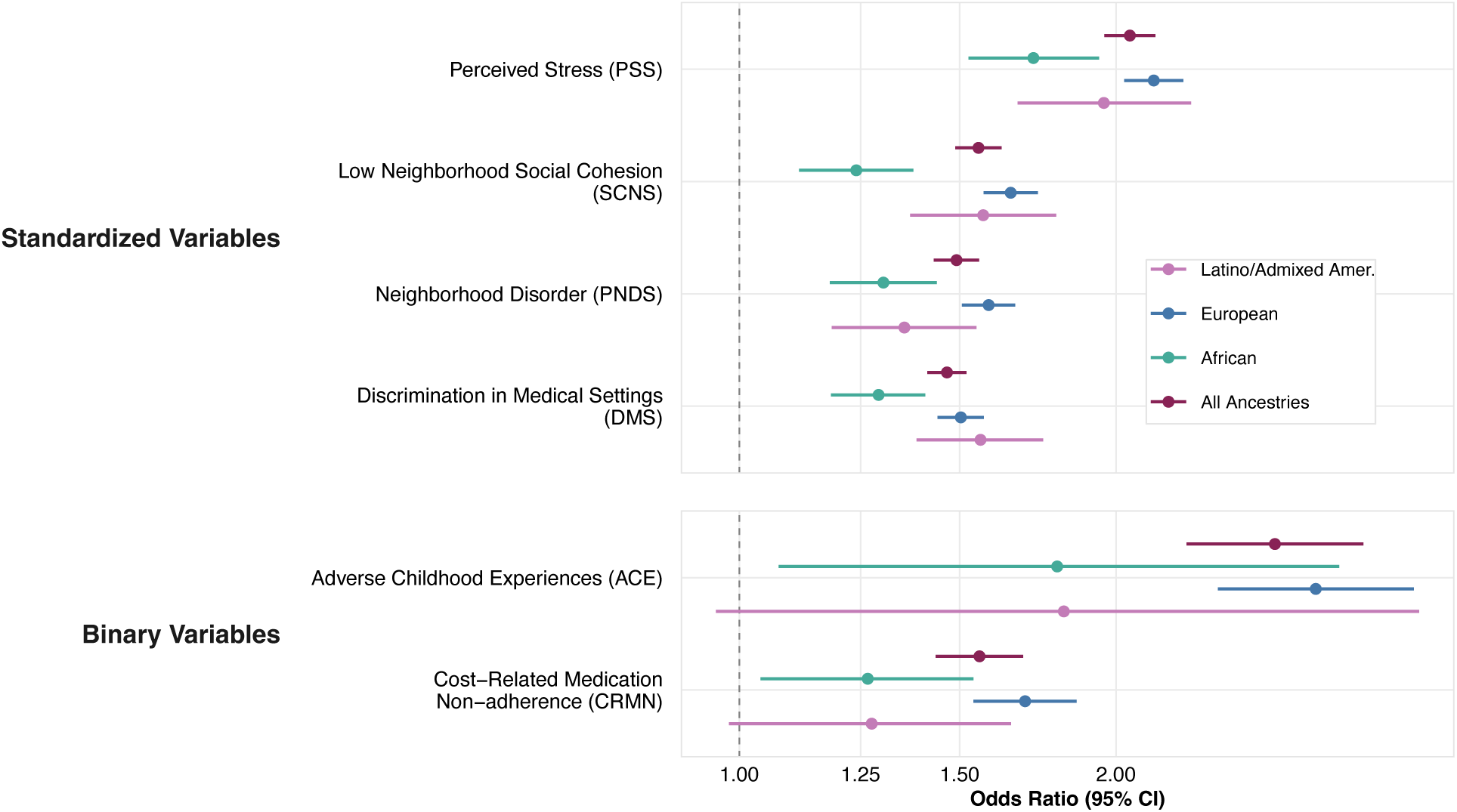
Association of social risk factors with bipolar disorder across ancestry groups. Forest plot displaying adjusted odds ratios (OR) and 95% confidence intervals (95% CI) for the association between social risk measures and bipolar disorder case status. Associations were estimated using logistic regression models adjusted for age, age^2^, sex/gender, EHR record length, record depth, and 16 genetic PCs. The upper panel displays results for continuous variables, which were Z-score standardized, such that ORs represent the risk associated with a one standard deviation increase in the social score. The lower panel displays results for binary indicators.

This pattern was largely mirrored for the examined binary indicators (CRMN and ACE). Reporting 4 or more ACEs showed the largest effect size of the binary variables tested, particularly in the EUR group (OR = 2.89; 95% CI 2.41-3.46; P = 7.54 × 10^-31^). Note that binary ORs (ACE, CRMN) represent the odds associated with exceeding a threshold, whereas continuous ORs (PSS, DMS, SCNS, PNDS) represent the odds per standard deviation increase; thus, these effects sizes are not directly comparable. While effect sizes were directionally consistent in the Latino/Admixed American group, the associations for ACE (OR = 1.82; 95% CI 0.96-3.49; P = 0.069) and CRMN (OR = 1.28; 95% CI 0.98-1.65; P = 0.065) did not reach statistical significance. Notably, effect sizes for social risk variables were consistently lower in the African ancestry group compared to the European and Admixed American groups. For example, the PSS association in the African ancestry group (OR = 1.72; 95% CI 1.52–1.94) was attenuated relative to the European group (OR = 2.14; 95% CI 2.03–2.26), a pattern observed across all six measures. Detailed association results by ancestry are provided in Table S4.

Beyond individual social-risk factors, we examined the cumulative impact of various risk factors, revealing a pattern wherein participants with a higher burden of social risk factors showed progressively higher prevalence of BD. (Figure S7).

### Combined Contributions of Polygenic and Social Risk to Bipolar Disorder

To examine the combined contributions of polygenic and social risk, we examined BD prevalence across PRS_BD_ deciles stratified by social risk group (Figure 4). Prevalence curves for the remaining four social variables are shown in Figure S8. Here, we highlight SCNS and CRMN, one continuously modeled and one binary-modeled variable, as these yielded the strongest interaction estimates and interaction significance values among the six tested variables. Notably, SCNS and CRMN also represent our two largest analytic sub-samples among the tested social risk variables (N = 49,478 and N = 64,009, respectively), affording greater statistical power to detect subtle interaction effects. For both Neighborhood Social Cohesion (SCNS; Figure 4A) and Cost-related Medication Nonadherence (CRMN; Figure 4B), individuals with high social risk (red lines) exhibited higher disease prevalence across all genetic risk deciles compared to those with low social risk (blue lines). Notably, individuals in the lowest genetic risk decile with high social burden often had disease prevalence rates comparable to or exceeding those in the highest genetic risk decile who had low social burden, underscoring the magnitude of social risk contributions. This contrast is further illustrated by direct comparison of BD prevalence between extreme combined risk groups (Figure S9). PRS_BD_ associations stratified by social risk level for all six variables are shown in Figure S10.

**Figure 4.**
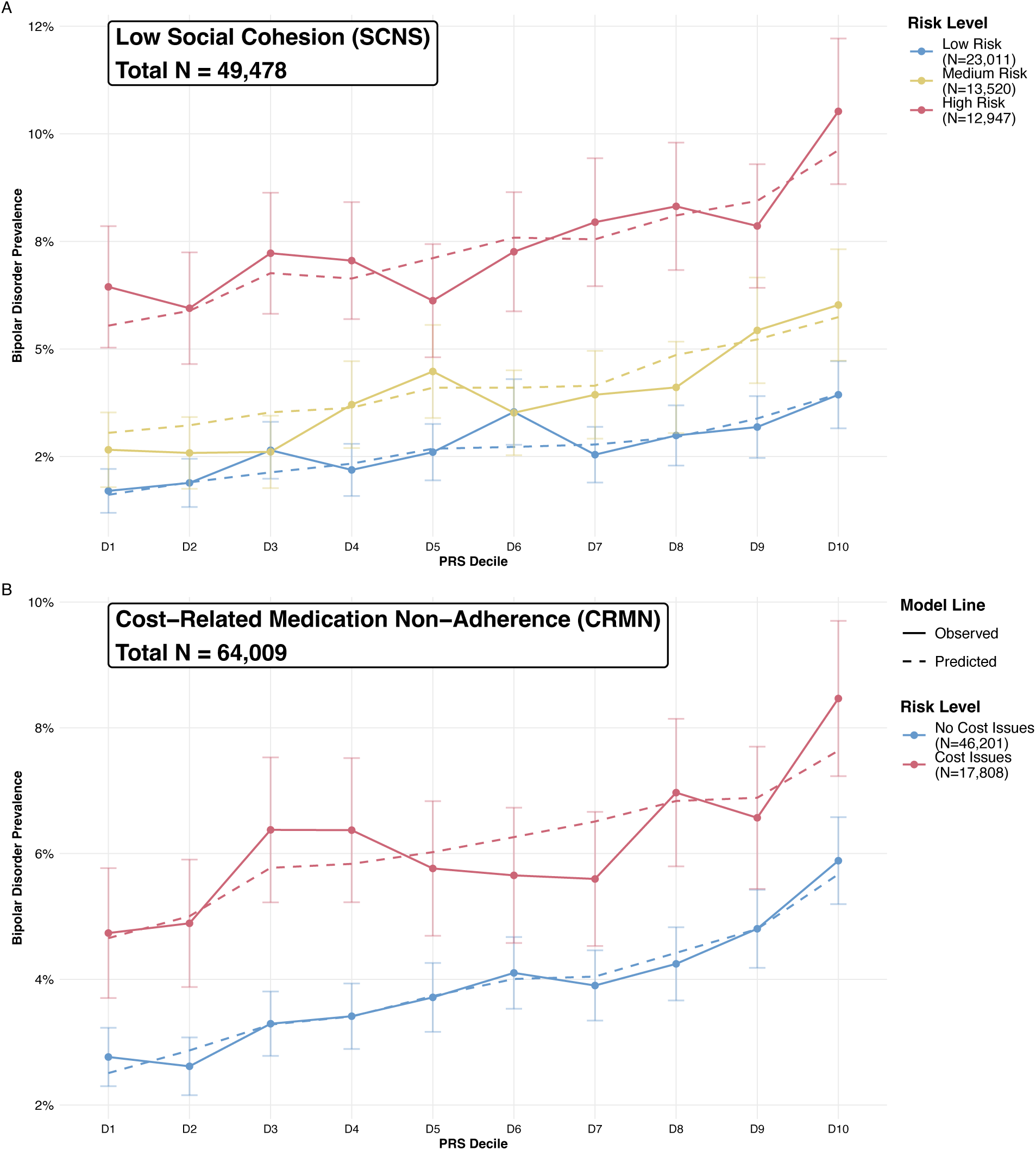
Polygenic Risk for Bipolar Disorder Stratified by Social Risk. Predicted (dashed) and observed (solid) prevalence of bipolar disorder (BD) across PRS deciles, stratified by social risk strata for (A) Low Neighborhood Social Cohesion (SCNS; tertiles) and (B) Cost-Related Medication Non-Adherence (CRMN; any vs. none). Predicted values are derived from logistic regression models adjusted for age, age², sex/gender, EHR record length, record depth, and 16 genetic PCs. PRS deciles were calculated within each analytic subsample. Error bars represent 95% confidence intervals. Sample sizes for each stratum are shown in the legend.

As has been discussed in the context of polygenic scores, coefficient estimates in logistic models can be biased when relevant predictors are omitted, even in the absence of true interactions.^35,79^ We therefore fit models for case status that included social risk, polygenic risk score (PRS), and their interaction as predictors. Across the six investigated factors, the interaction terms were generally small in magnitude, with 95% confidence intervals often crossing or approaching zero (Figure 4; Table 3). Both SCNS and CRMN showed nominally significant interactions.

**Table 3.** Interaction Between Polygenic Risk Score and Social Risk Factors.

| Variable | N | N_C<br>ases | PRS OR (95%<br>CI) | SDoH OR (95%<br>CI) | Interaction $\beta$<br>(95% CI) | P-value | FDR |
| --- | --- | --- | --- | --- | --- | --- | --- |
| Perceived Stress (PSS) | 48881 | 2086 | 1.20 (1.15–1.26) | 2.04 (1.95–2.14) | -0.043 (-0.087–<br>0.001) | 0.055 | 0.096 |
| Discrimination in<br>Medical Settings (DMS) | 49402 | 2113 | 1.22 (1.17–1.27) | 1.46 (1.41–1.52) | 0.007 (-0.028–<br>0.042) | 0.692 | 0.692 |
| Cost-Related<br>Medication Non-Adherence (CRMN) | 64009 | 2883 | 1.23 (1.18–1.27) | 1.54 (1.42–1.67) | -0.085 (-0.164–<br>-0.006) | 0.034 | 0.096 |
| Low Social Cohesion<br>(SCNS) | 49478 | 2120 | 1.21 (1.16–1.27) | 1.55 (1.48–1.62) | -0.050 (-0.091–<br>-0.010) | 0.016 | 0.093 |
| Perceived<br>Neighborhood Disorder<br>(PNDS) | 48900 | 2077 | 1.22 (1.17–1.28) | 1.49 (1.43–1.55) | -0.031 (-0.070–<br>0.008) | 0.120 | 0.144 |
| Adverse Childhood<br>Experiences (ACE) | 17098 | 700 | 1.23 (1.14–1.33) | 2.64 (2.24–3.11) | -0.146 (-0.301–<br>0.009) | 0.064 | 0.096 |

Logistic regression models adjusted for age, age<sup>2</sup>, sex/gender, 16 genetic PCs, EHR record length, and record depth. Interaction $\beta$ represents the coefficient of the PRS × Social Risk product term on the log-odds scale. Negative values indicate sub-multiplicative interaction. FDR: Benjamini-Hochberg adjusted p-values across six tests.

For SCNS, we observed a nominally significant interaction between social risk category and PRS_BD_ (N = 49,478; P*int* = 0.016). Notably, the effect of social risk (OR = 1.55) was larger than the effect of genetic risk (OR = 1.21), and the negative estimate of the interaction coefficient (-0.05) indicated that a slightly attenuated association with genetic risk was observed in high social risk group. We observed a similar pattern for CRMN (N = 64,009; P = 0.034), where the social risk effect (OR = 1.54) exceeded the genetic risk effect (OR = 1.23), and the negative interaction coefficient (-0.09) again suggested a slightly attenuated association with genetic risk in the high social risk group. However, these interaction terms did not remain significant after correction for multiple testing (Benjamini-Hochberg FDR > 0.05). Interaction estimates were broadly consistent when re-estimated under alternative case definition strategies (Figure S11). Notably, across all six interaction models, the odds ratio for the social risk term (range: OR = 1.47–2.05 for continuous measures; OR = 1.54–2.89 for binary measures) exceeded that of the PRS (range: OR = 1.19–1.23), reinforcing that social risk factors contributed substantially to BD liability beyond what was captured by polygenic risk alone.

## Discussion

In this study, we evaluated the contributions of polygenic risk scores (PRS) and social risk factors to bipolar disorder (BD) case status in the All of Us Research Program (AoU). We successfully reproduced the association between PRS and BD case status across all ancestries, though with low and variable predictive performance. We also evaluated the effect of several social risk factors on bipolar disorder liability: perceived stress (PSS), experiences of discrimination in medical settings (DMS), perceived neighborhood disorder (PNDS), neighborhood social cohesion (SCNS), non-adherence to prescribed medication due to cost barriers (CRMN), and reports of adverse childhood experiences (ACE). Although these associations were estimated in a non-random sample with complex missingness structures, limiting our ability to interpret population-level effects, the observed effect sizes were substantial and often eclipsed those of genetic risk. While we hypothesized that social risk might make those with the highest polygenic risk even more susceptible to development of BD, our results did not yield statistically significant interaction terms after correction. Instead, we observed primarily additive contributions of genetic and social risk. However, these patterns should be interpreted with caution given the attenuated PRS performance and modest sample sizes available for interaction testing.

Consistent with recent epidemiological work, our results underscore the sizable role of social and environmental risk factors in psychiatric outcomes. The effect sizes we observed for social variables were substantial; for instance, reporting four or more adverse childhood experiences (ACEs) was associated with a nearly 3-fold increase in odds of BD in European ancestries (OR=2.89; 95% CI 2.41-3.46). Similarly, Perceived Stress (PSS) showed a strong linear association across all groups, with each standard deviation increase in PSS associated with an approximately 2-fold increase in BD odds (OR = 2.05; 95% CI 1.96-2.15 in the All Ancestries analysis). However, because the AoU cohort is not a random population sample, containing unmodelled patterns of missingness in EHR and survey data, these effect sizes should be interpreted as associations within this specific analytic sample rather than as direct estimates of population-level risk.^80^

Among the social risk variables tested, ACE is unique in that it captures retrospective reports of childhood experiences that typically precede adult BD onset. This aspect of temporality supports a somewhat more directional interpretation than is possible for the other measures, but it is important to note that a causal interpretation cannot be assumed. Still, the nearly 3-fold OR observed for ACE in the European ancestry group is consistent with a growing body of literature linking childhood maltreatment to earlier BD onset and greater illness severity.^21,26^

A notable pattern observed in our ancestry-stratified analyses was that effect sizes for social risk variables were consistently lower in the African ancestry group compared to the European and Latino/Admixed American groups. Several factors may contribute to this effect. First, differential rates of misdiagnosis in Black Americans with BD (who are more likely to be misdiagnosed with schizophrenia)^27,28^ may attenuate differences between cases and controls in this group by including true BD cases among controls. Second, differential survey response patterns and the healthy volunteer bias inherent in AoU may operate differently across ancestry groups, as indicated by our comparison of survey completion rates across ancestries and case/control groups (Figure S3). These findings warrant careful interpretation and underscore the need to investigate how structural factors shape both diagnostic accuracy and the measurement of social risk in diverse cohorts.

When modeling combined contributions of PRS and social risk factors, we observed that social risk consistently contributed substantial additional explanatory power beyond genetic risk alone. In several models, individuals in the lowest genetic risk decile with high social burden exhibited BD prevalence rates comparable to or exceeding those in the highest genetic risk decile with low social burden, underscoring the magnitude of social risk contributions. Across the six investigated social risk factors, interaction terms between PRS and social risk were generally small in magnitude and did not survive multiple testing correction (FDR > 0.05). The nominally significant negative interaction coefficients observed for SCNS and CRMN suggest a pattern in which the association between genetic risk and BD is slightly attenuated in high-social-risk groups. One interpretation is that when social risk is sufficiently high, baseline BD prevalence is already elevated, and the incremental contribution of genetic liability is proportionally smaller. Given the attenuated PRS performance in our cohort and the relatively modest sample sizes available for interaction testing, larger studies will be needed to further assess whether polygenic risk and social risk interact in their association with BD.

A notable finding in our study was the attenuation of PRS predictive power compared to prior literature. While the latest PGC meta-analysis for BD reported R^2^ liability-scale estimates of 9% in clinical cohorts, our analysis yielded estimates of just 1.86% in European ancestries. This discrepancy likely reflects the “phenotyping gap” observed between curated research cohorts and real-world data. Discovery GWAS typically rely on strict, structured clinical interviews to define cases, whereas our study relied on EHR diagnostic codes and medication records. Psychiatric disorders are prone to high rates of diagnostic flux and misclassification – estimates suggest up to 40% of BD patients are initially misdiagnosed, with the rate and nature of misdiagnosis varying by racial group.^28,81^ The noise inherent in EHR-based phenotyping inevitably dilutes the genetic signal. In addition to random noise, recent simulations demonstrate that diagnostic inaccuracies based on EHR codes and self-report instruments introduce heritable confounders that generate systematic off-target genetic associations which cannot be overcome by increasing sample size alone.^50^ The extent to which such confounders operate differently across ancestry groups — for instance through differential patterns of healthcare utilization, diagnostic practices, or case ascertainment — remains an open question that may further contribute to the ancestry-stratified differences in both PRS performance and social risk effect sizes observed in our study. Consistent with this, the PGC BD GWAS found that SNP-heritability differed markedly by ascertainment method (h^2^SNP_clinical_ = 0.22; h^2^SNP_EHR_ = 0.05; h^2^SNP_self-report_ = 0.08), and that genetic correlations between clinical and self-report BD cohorts were well below unity (rG = 0.47), indicating partially distinct genetic architectures.^12,50^ Finally, PRS derived from shallow phenotypes have been shown to predict many phenotypes beyond the target disorder, suggesting that higher R^2^ values in curated research cohorts may partly reflect nonspecific genetic signals. Our results may therefore provide a more realistic estimate of how current PRS perform when applied to the heterogeneous clinical reality of general healthcare systems. Taken together, these findings reinforce the importance of deep, specific phenotyping in psychiatric genomics, not only for GWAS discovery, but also for downstream applications and interpretation of PRS in clinical and research settings.

Methodological choices in PRS application may also contribute to the observed attenuated performance. Although we used the O’Connell multi-ancestry GWAS as the discovery sample for PRS calculation, we chose to apply a European-ancestry LD reference panel, as did O’Connell et al., given the high proportion (∼82% by effective sample size) of European-ancestry participants in the source GWAS. Methods that better account for complex admixture or are designed for diverse cohorts could improve accuracy. However, with both the GWAS discovery sample and the application cohort being majority European, this effect is likely modest. Additionally, the impact of case ascertainment on PRS performance varies by ancestry in ways that may complicate cross-population comparisons. O’Connell et al. found that excluding self-reported cases improved PRS performance in European and East Asian target cohorts, but in an African-ancestry target cohort, inclusion of self-reported cases substantially increased variance explained.^12^ This suggests that optimal strategies for identifying genetic signals may differ by ancestry group, potentially reflecting differences in how the phenotyping biases described above, case ascertainment practices, and linkage disequilibrium structure interact across populations, underscoring the need for ancestry-aware PRS pipelines in diverse biobank settings.

Our study further highlights the persistent “portability gap” seen in complex trait genetics. Consistent with the well-documented Eurocentric bias in GWAS discovery, PRS discriminative ability was highest in European and Admixed American groups and markedly lower in the African ancestry group. In our study, lower PRS performance in the African ancestry group likely reflects a combination of this portability gap and the ancestry-specific phenotyping and ascertainment biases described above, though disentangling these contributions remains challenging. We were also unable to robustly assess PRS performance in East Asian, South Asian, and Middle Eastern ancestry groups, due to low sample sizes in these groups. This discrepancy in predictive power raises equity concerns: until large-scale genomic datasets prioritize the recruitment of non-European populations, the clinical implementation of PRS risks exacerbating existing community health disparities. Thankfully, there are many groups working to expand genetic diversity and equity to non-European populations, including the Latin American Genomics Consortium, the Ancestral Populations Network, the Nigerian 100K Genome Project, and the Sangre por Salud Biobank, among others.^82–87^ Crucially, these efforts must be conducted in partnership with represented communities and with attention to historical harms and the ethical obligations of genomic research in underrepresented populations, prioritizing community trust, partnership, and safety.^88–91^

We acknowledge several limitations of the present study. Our cohort of 7,000 BD cases, though substantial for PRS validation, may lack the statistical power required to detect subtle GxE interactions, particularly given the attenuated variance explained by PRS in our EHR-derived cohort. In the All Ancestries group, we did observe nominal negative interaction coefficients for several social risk factors, a pattern warranting further study in larger samples. The AoU cohort is subject to “healthy volunteer” bias, with participants tending to be healthier, wealthier, and more technologically literate than the general U.S. population, potentially attenuating the range of social adversity observed.^80,92^ This bias is likely compounded in our analyses, which further restrict to the subset of participants who completed optional SDOH surveys – a group which is likely even less representative of the broader population.^80^ With respect to phenotyping, our EHR-based phenotyping strategy, while designed to balance sensitivity and specificity, does not capture BD subtype information, nor does it account for the episodic and cyclical nature of the disorder. Stricter and/or temporal case definitions might improve specificity but would further reduce the already limited samples sizes, particularly in non-European ancestry groups.

Our results highlight the importance of holistic models of psychiatric epidemiology which integrate genetic risk with the social and environmental context that ultimately shapes disease manifestation. The consistent and large effect sizes observed for social risk factors, which often exceeded those of genetic risk, suggest that precision psychiatry frameworks that fail to include social or environmental risk information provide an incomplete picture of individual-level risk. Importantly, many of the social risk factors examined here are modifiable or addressable through targeted or community-level interventions, unlike genetic liability. Improved risk prediction through joint genetic and social models has direct clinical relevance: earlier and more accurate identification of at-risk individuals can facilitate timely intervention, which has been associated with lower rates of suicide and psychiatric comorbidities in BD. Future work should expand these analyses to larger and more diverse cohorts, incorporate additional modifiable risk factors, and evaluate whether combined genetic-social risk models improve clinical outcomes.

Together, these findings support continued development of risk prediction frameworks that integrate genetic liability with social context for more accurate and equitable risk prediction.

## Data Availability

This study used data from the All of Us Research Programs Controlled Tier Dataset version v8, available to authorized users on the Researcher Workbench. All analyses were done using Jupyter Notebooks and R on the All of Us workbench. Following controlled access registration and training, we will make our notebooks available upon request through the All of Us workbench.

## Acknowledgments

We gratefully acknowledge All of Us participants for their contributions, without whom this research would not have been possible. We also thank the National Institutes of Health’s All of Us Research Program for making available the participant data examined in this study. This material is based upon work supported by the National Science Foundation Graduate Research Fellowship Program under Grant No. DGE-2439854 awarded to R.R.S. Any opinions, findings, and conclusions or recommendations expressed in this material are those of the author(s) and do not necessarily reflect the views of the National Science Foundation, or the National Institutes of Health.

## Author contributions

R.R.S. conceived the study, curated and analyzed the data, secured funding, created all figures, and wrote the manuscript. M.H., R.G.M., L.G., and L.M.R. provided relevant expertise on the study design and reviewed the manuscript. M.I.L. co-conceived the study, supervised the work, and reviewed the manuscript.

## Web resources

All analyses were performed on the All of Us workbench, which uses Google Cloud Infrastructure. (https://www.researchallofus.org/data-tools/workbench/).

## Data and code availability

This study used data from the *All of Us* Research Program’s Controlled Tier Dataset version v8, available to authorized users on the Researcher Workbench. All analyses were done using Jupyter Notebooks and R on the *All of Us* workbench. Following controlled access registration and training, we will make our notebooks available upon request through the *All of Us* workbench.

## Declaration of generative AI and AI-assisted technologies in the writing process

During the preparation of this work the authors used Claude to organize ideas, refactor code, and format the manuscript. The authors reviewed and edited all AI-generated content and take full responsibility for the content of the publication.

