## Supplemental Information for "Considering social risk alongside genetic risk for bipolar disorder in the All of Us Research Program"

### Supplemental material and methods

#### Diagnostic codes for BD case definition

##### ICD-10-CM codes

- F30.1* (manic episode without psychotic symptoms)
- F30.2 (manic episode, severe, with psychotic symptoms)
- F30.3 (manic episode in partial remission)
- F30.4 (manic episode in full remission)
- F30.8 (other manic episodes)
- F31.0 (bipolar disorder, current episode hypomanic)
- F31.1* (bipolar disorder, current episode manic without psychotic features)
- F31.2 (bipolar disorder, current episode manic, severe with psychotic features)
- F31.3* (bipolar disorder, current episode depressed, mild or moderate severity)
- F31.4 (bipolar disorder, current episode depressed, severe, without psychotic features)
- F31.5 (bipolar disorder, current episode depressed, severe, with psychotic features)
- F31.6* (bipolar disorder, current episode mixed)
- F31.7* (bipolar disorder, currently in remission)
- F31.8* (other bipolar disorders)
- F31.9 (bipolar disorder, unspecified)

##### ICD-9-CM codes

- 296.0* (bipolar I disorder, single manic episode)
- 296.1* (manic disorder, recurrent episode)

- 296.4* (bipolar I disorder, most recent episode manic)
- 296.5* (bipolar I disorder, most recent episode depressed)
- 296.6* (bipolar I disorder, most recent episode mixed)
- 296.7 (bipolar I disorder, most recent episode unspecified)
- 296.8* (other and unspecified bipolar disorders)

** Asterisk denotes that all sub-codes within the code family were included.*

#### Medications for BD case definition

- Lithium
  - Lithium carbonate
  - Lithium citrate
  - Lithium chloride
  - Lithium sulfate
- Valproate
- Carbamazepine
- Lamotrigine

#### Exclusion codes for case definition

- Psychotic disorder (OMOP Concept ID: 436073)
- Schizophrenia (OMOP Concept ID: 435783)
- Paranoid schizophrenia (OMOP Concept ID: 433450)
- Chronic schizophrenia (OMOP Concept ID: 435782)
- Schizoaffective schizophrenia (OMOP Concept ID: 432597)
- Chronic schizoaffective schizophrenia (OMOP Concept ID: 441835)
- Chronic paranoid schizophrenia (OMOP Concept ID: 436944)

- Undifferentiated schizophrenia (OMOP Concept ID: 4008566)
- Simple schizophrenia (OMOP Concept ID: 436067)
- Disorganized schizophrenia (OMOP Concept ID: 441828)
- Schizoaffective disorder (OMOP Concept ID: 4286201)
- Organic mood disorder (OMOP Concept ID: 373176)
- Major depressive disorder (OMOP Concept ID: 4152280)

##### Exclusion codes for control definition

- 291* (alcohol-induced mental disorders)
- 292* (drug-induced mental disorders)
- 295* (schizophrenic disorders)
- 296* (episodic mood disorders)
- 297* (paranoid states, delusional disorders)
- 298* (other nonorganic psychoses)
- 301* (personality disorders)

### Supplemental figure legends

**Figure S1. PRS Density Distributions by Ancestry Before and After PC Adjustment**

Density plots of standardized $PRS_{BD}$ (SD units) for each genetically predicted ancestry group (African, European, Latino/Admixed American). Top panel: unadjusted PRS (raw scores standardized globally). Bottom panel: PC-adjusted PRS (residualized on 16 genetic PCs, then standardized globally).

**Figure S2. Social Risk Data Quality**

(A) UpSet plot showing patterns of item-level data availability across the six social risk surveys. Horizontal bars show total participants with data for each survey; vertical bars show intersection sizes. (B) Comparison of score distributions when using imputed data (red) versus complete cases only (blue) for each social risk measure. (C) Pairwise Pearson correlations among the six social risk measures.

**Figure S3. Survey Completion Rates by Ancestry and Case Status**

Proportion of participants completing each social risk survey, stratified by genetically predicted ancestry group (African, European, Latino/Admixed American) and EHR-defined case/control status.

**Figure S4. Self-Report Concordance with EHR Phenotyping**

(A) Estimated probability of endorsing each self-report indicator as a function of normalized PRS, with 95% confidence bands. Indicators: currently receiving BD medication, currently seeing a doctor for BD, having a parent with BD, and self-reported BD. (B) Proportion of EHR-defined cases and controls endorsing each self-report indicator.

**Figure S5. PRS Distributions by EHR Case Assignment Strategy**

Violin plots of normalized $PRS_{BD}$ distributions across three EHR-based case assignment methods: 2+ diagnostic codes (most stringent), code + concordant medication, and high medication volume (Med++).

**Figure S6. Theoretical Absolute Risk of Bipolar Disorder by PRS**

Theoretical absolute risk curves as a function of standardized PRS, derived from liability-scale $R^2$ estimates assuming 2% population prevalence. Curves shown for All Ancestries, European,

African, and Latino/Admixed American groups. Dashed horizontal line indicates 2% population prevalence.

**Figure S7. Cumulative Social Risk Dose-Response**

Observed BD prevalence as a function of the number of high-risk social factors (0–6). High-risk was defined as top tertile for continuous measures and threshold exceedance for binary measures. Error bars represent 95% confidence intervals. Sample sizes annotated above each bar.

**Figure S8. BD Prevalence Across PRS Deciles Stratified by Social Risk**

Predicted (dashed) and observed (solid) prevalence of BD across PRS deciles, stratified by social risk level for (A) Perceived Stress, (B) Discrimination in Medical Settings, (C) Perceived Neighborhood Disorder, and (D) Adverse Childhood Experiences.

**Figure S9. BD Prevalence by Combined Genetic and Social Risk Group**

Observed BD prevalence across four combined risk groups defined by PRS level (top/bottom decile) and social risk level (high/low) for (A) Low Neighborhood Social Cohesion (SCNS) and (B) Cost-Related Medication Non-Adherence (CRMN). Error bars represent 95% CIs.

**Figure S10. PRS Association Stratified by Social Risk Level**

$PRS_{BD}$ odds ratios and 95% CIs estimated separately within social risk strata for all six social risk variables: (A) Perceived Stress, (B) Discrimination in Medical Settings, (C) Cost-Related Medication Non-Adherence, (D) Low Social Cohesion, (E) Perceived Neighborhood Disorder, (F) Adverse Childhood Experiences.

**Figure S11. Interaction Estimate Sensitivity to Case Definition**

Interaction coefficients (PRS × social risk) estimated under alternative EHR case definitions: 2+ diagnostic codes and code + concordant medication. Points show estimated interaction β; horizontal lines show 95% CIs. Dashed vertical lines at zero.

### Supplemental figures

**Figure S1. PRS Density Distributions by Ancestry and Case Status**

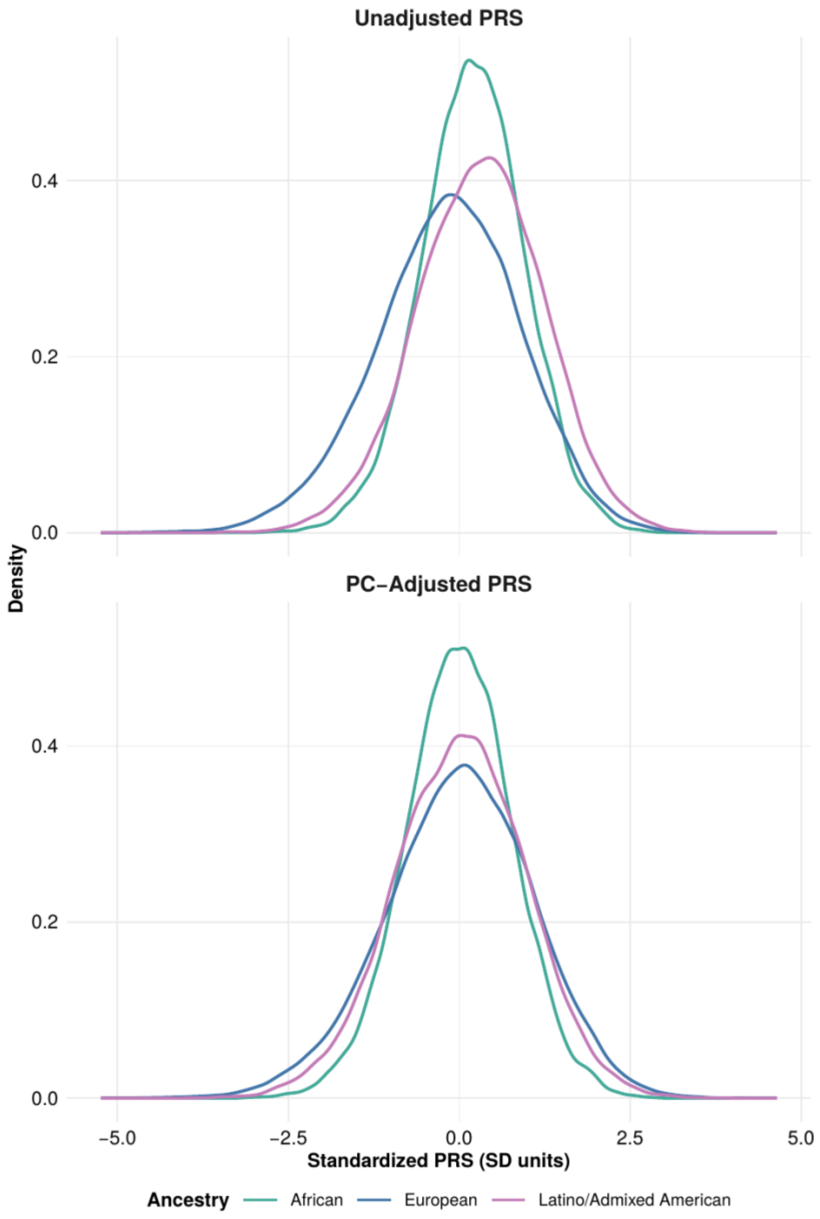

**Figure S2A. Item-Level Missingness Patterns**

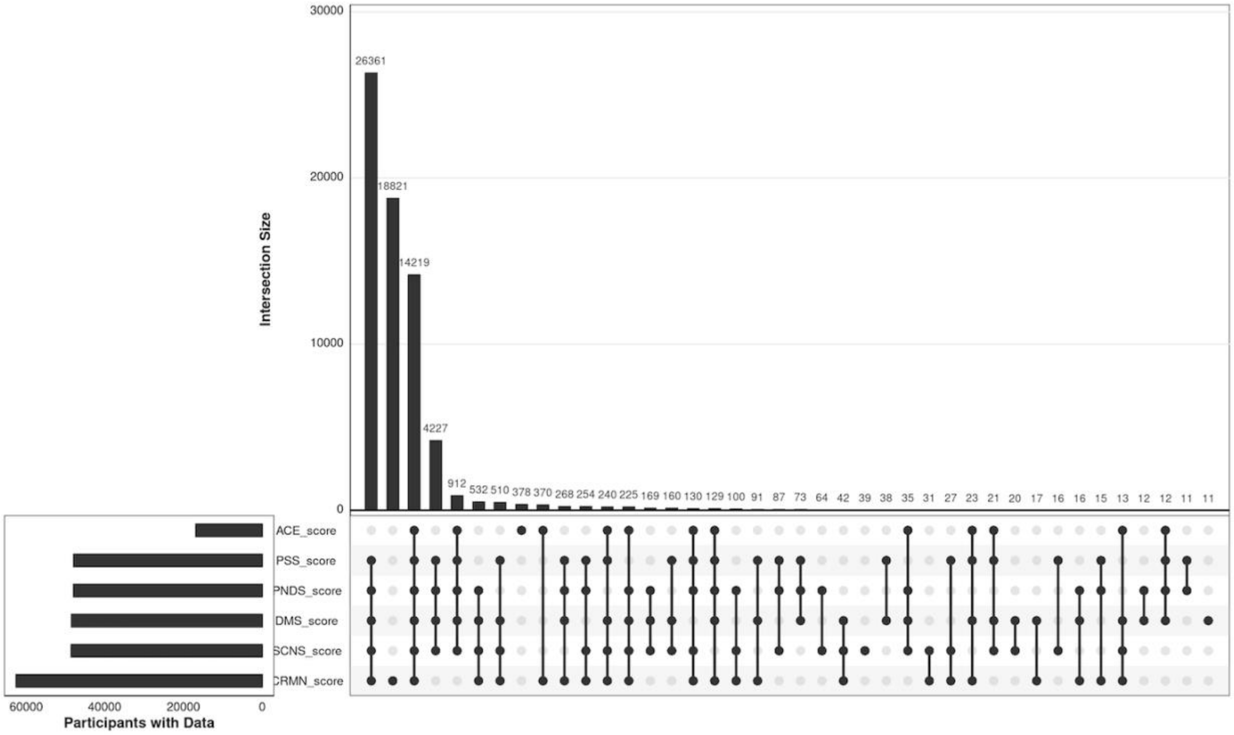

127 **Figure S2B. Imputed vs. Complete-Case Score Distributions**

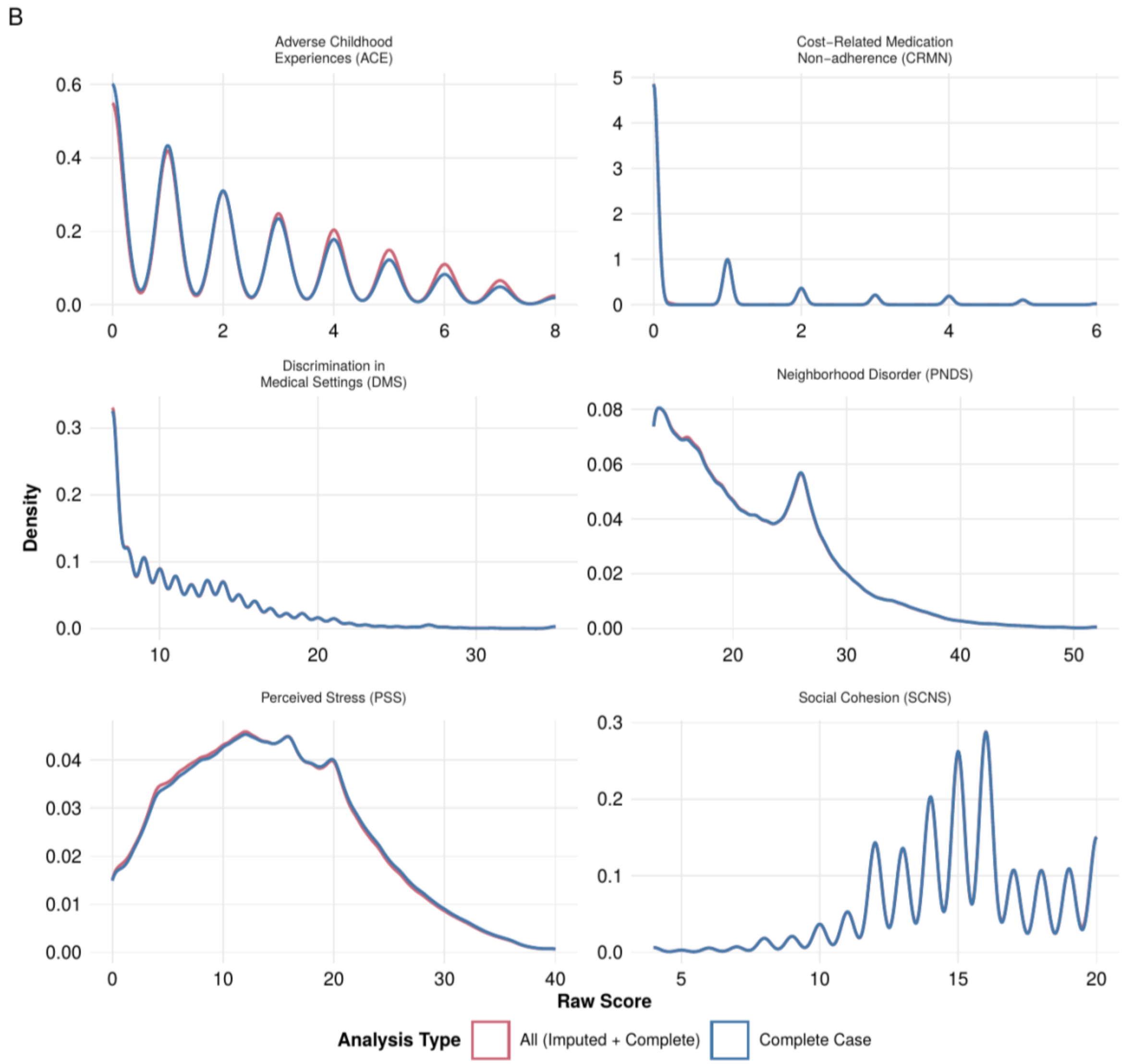


128

129

**Figure S2C. Inter-Scale Correlations**

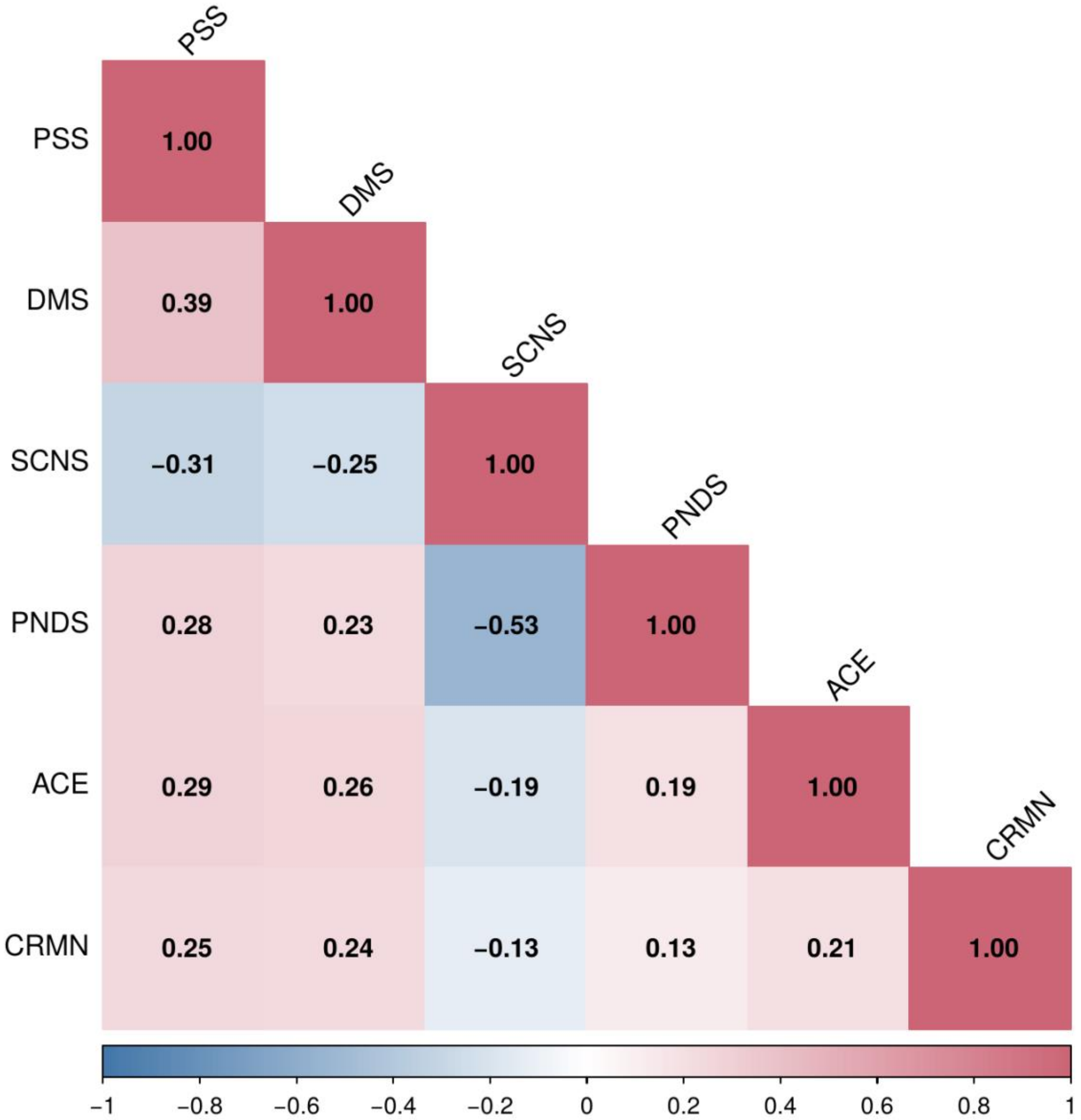

**Figure S3. Survey Completion Rates by Ancestry and Case Status**

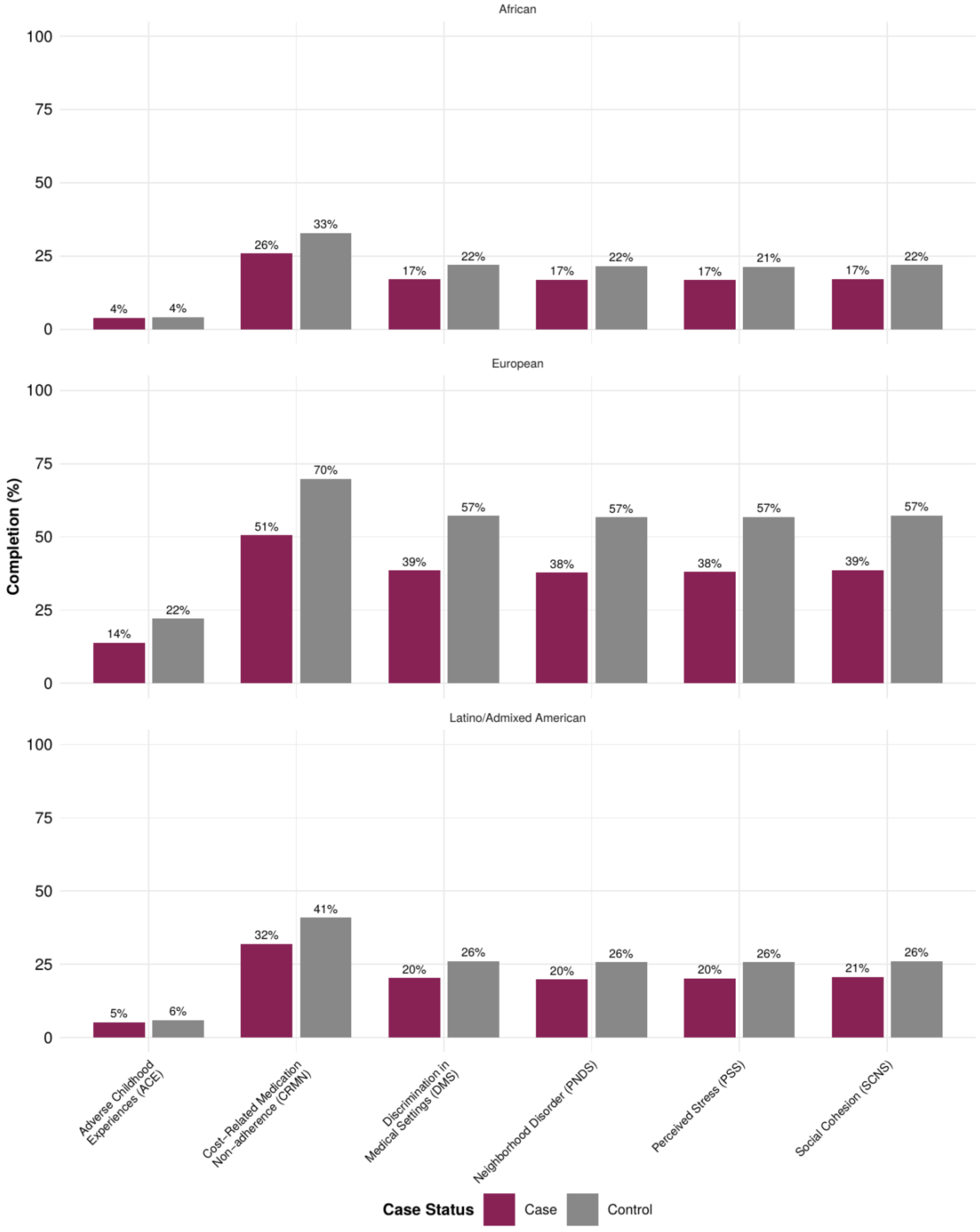

**Figure S4. Self-Report Concordance with EHR Phenotyping**

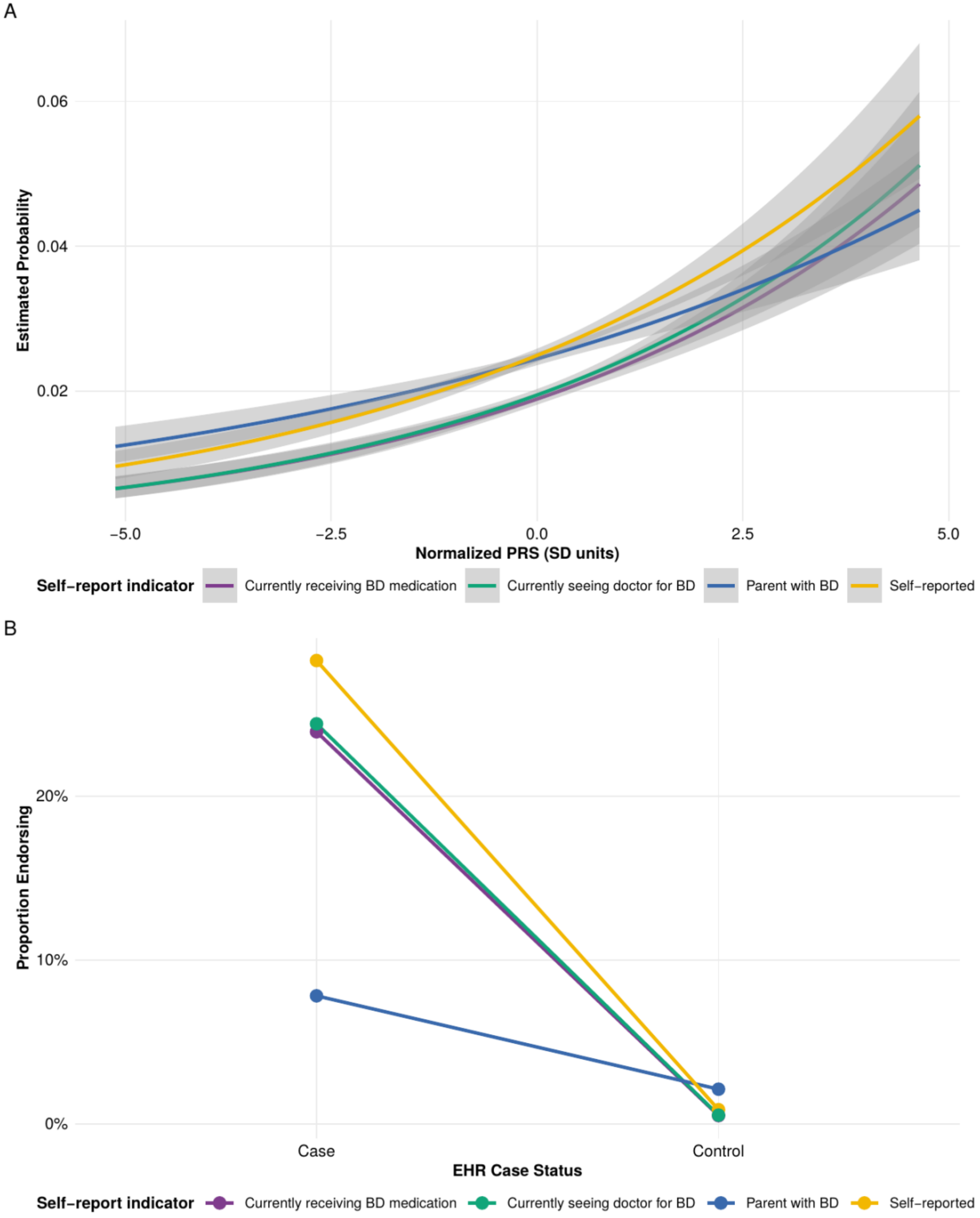

139 **Figure S5. PRS Distributions by EHR Case Assignment Strategy**

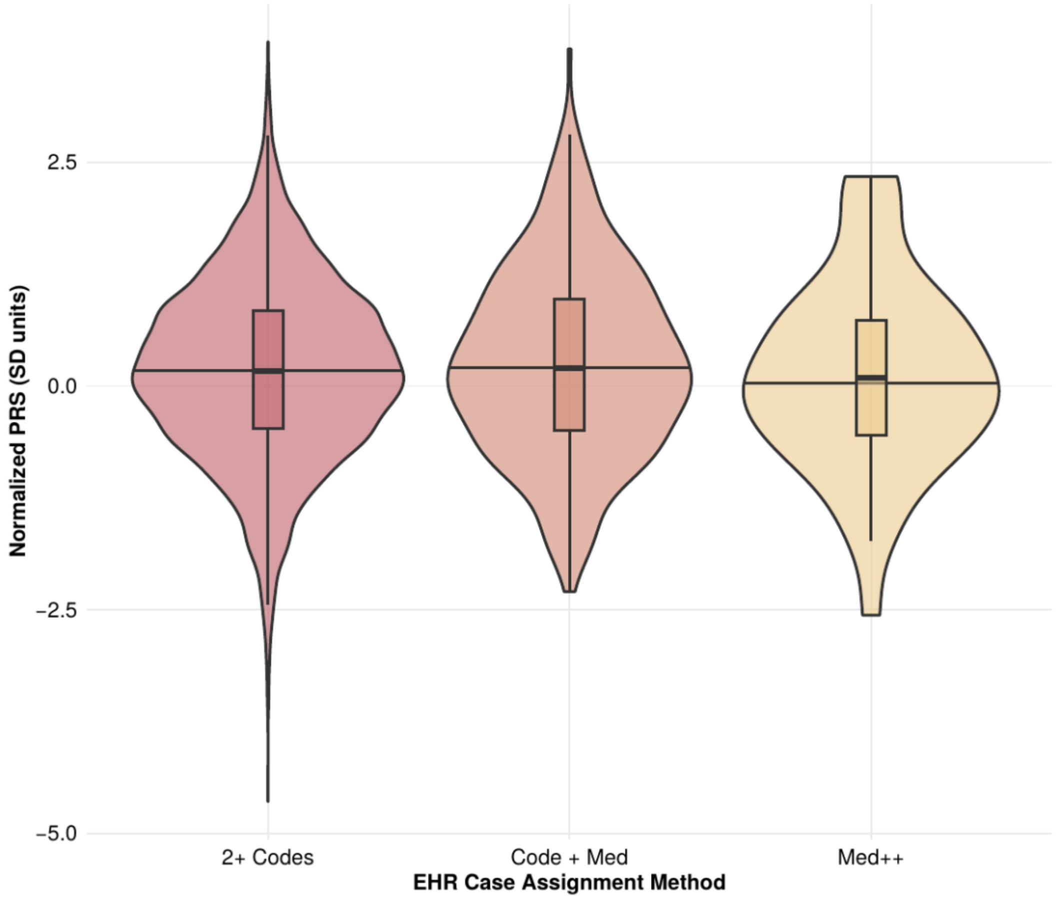


140

141

**Figure S6. Theoretical Absolute Risk of Bipolar Disorder by PRS**

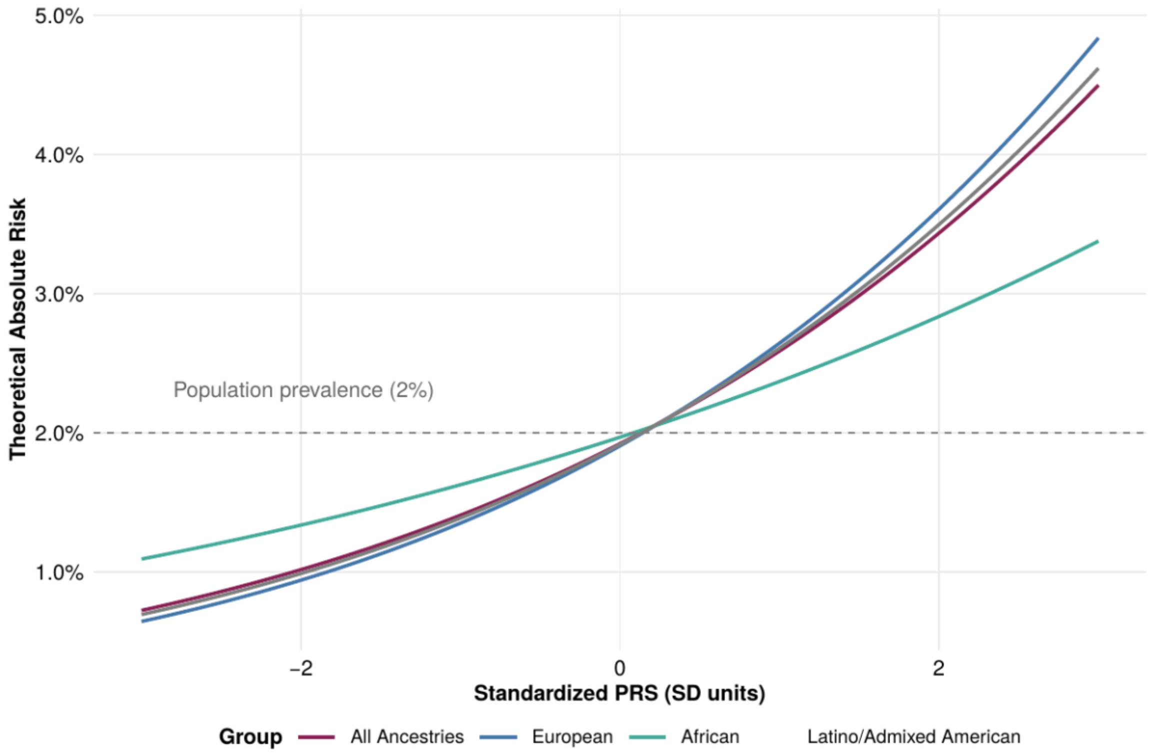


**Figure S7. Cumulative Social Risk Dose-Response**

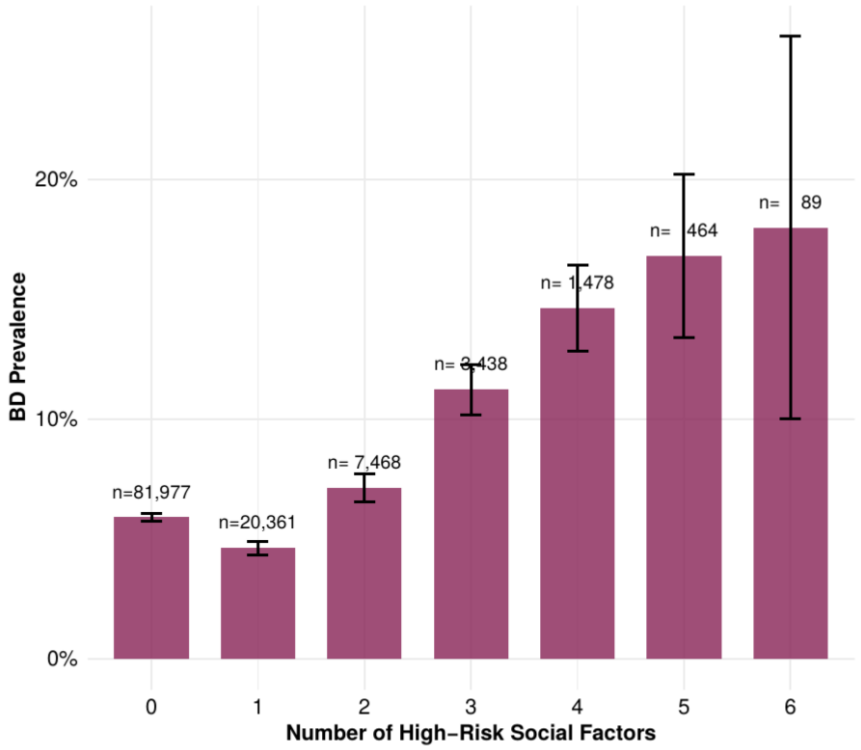

**Figure S8. BD Prevalence Across PRS Deciles (Remaining Variables)**

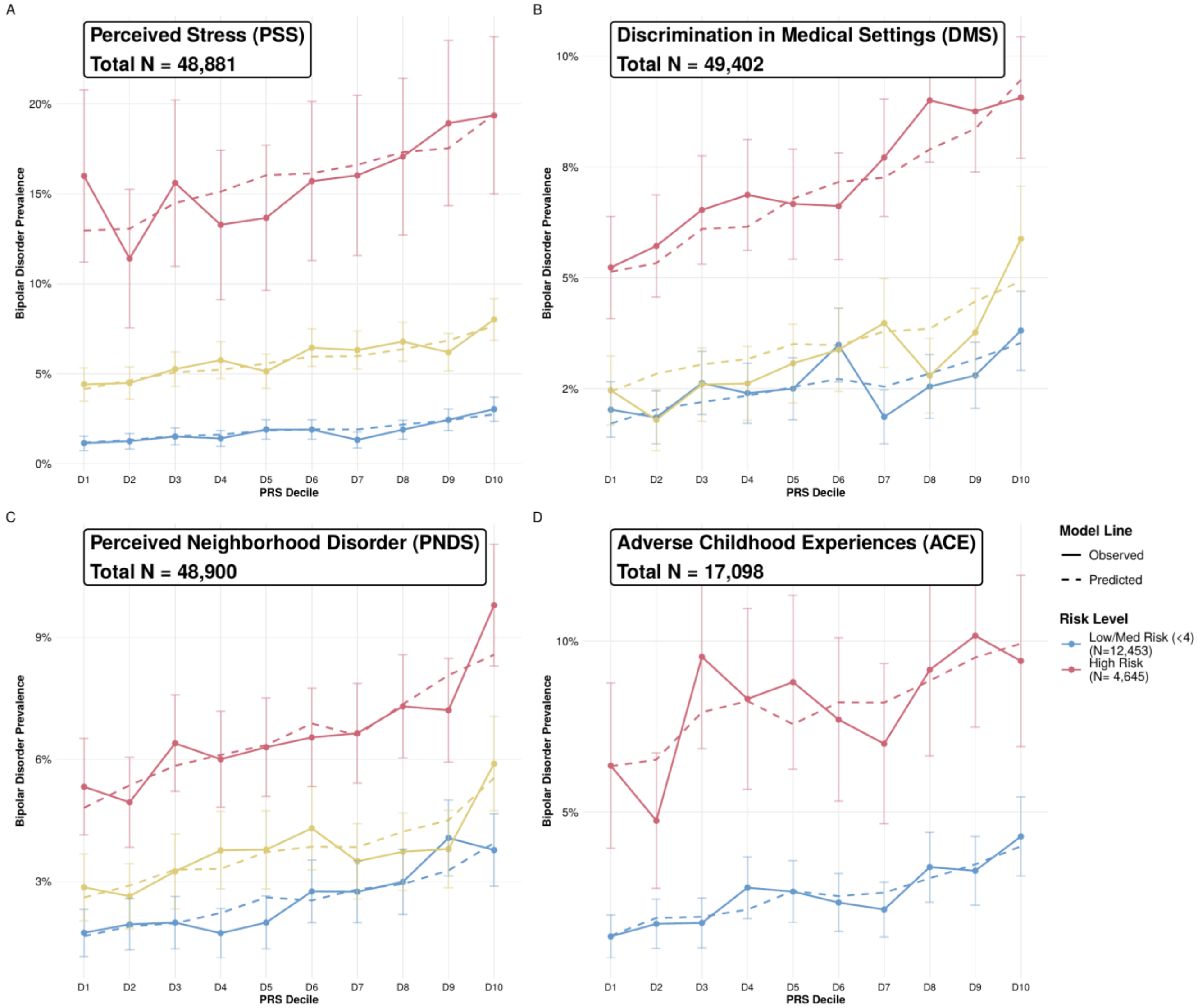

**Figure S9. BD Prevalence by Combined Genetic and Social Risk Group**

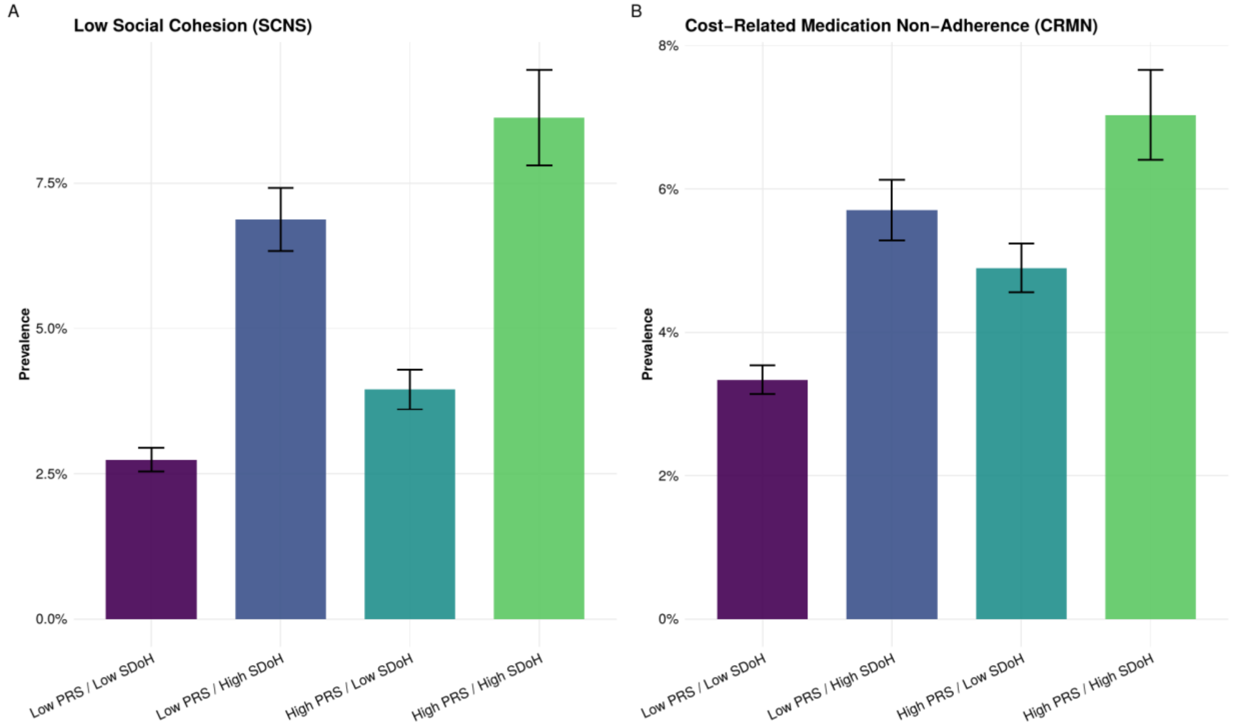

**Figure S10. PRS Association Stratified by Social Risk Level**

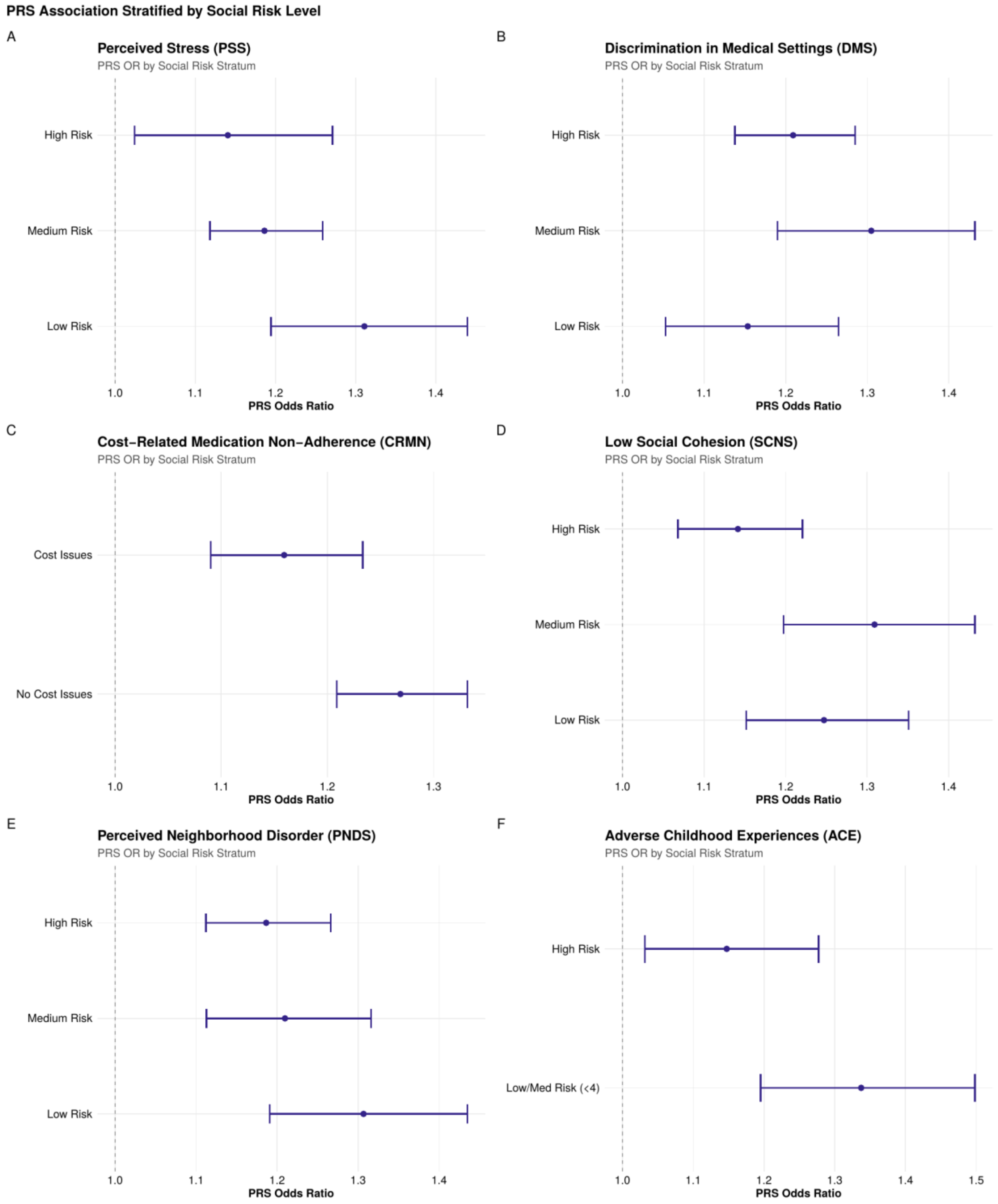

**Figure S11. Interaction Estimate Sensitivity to Case Definition**

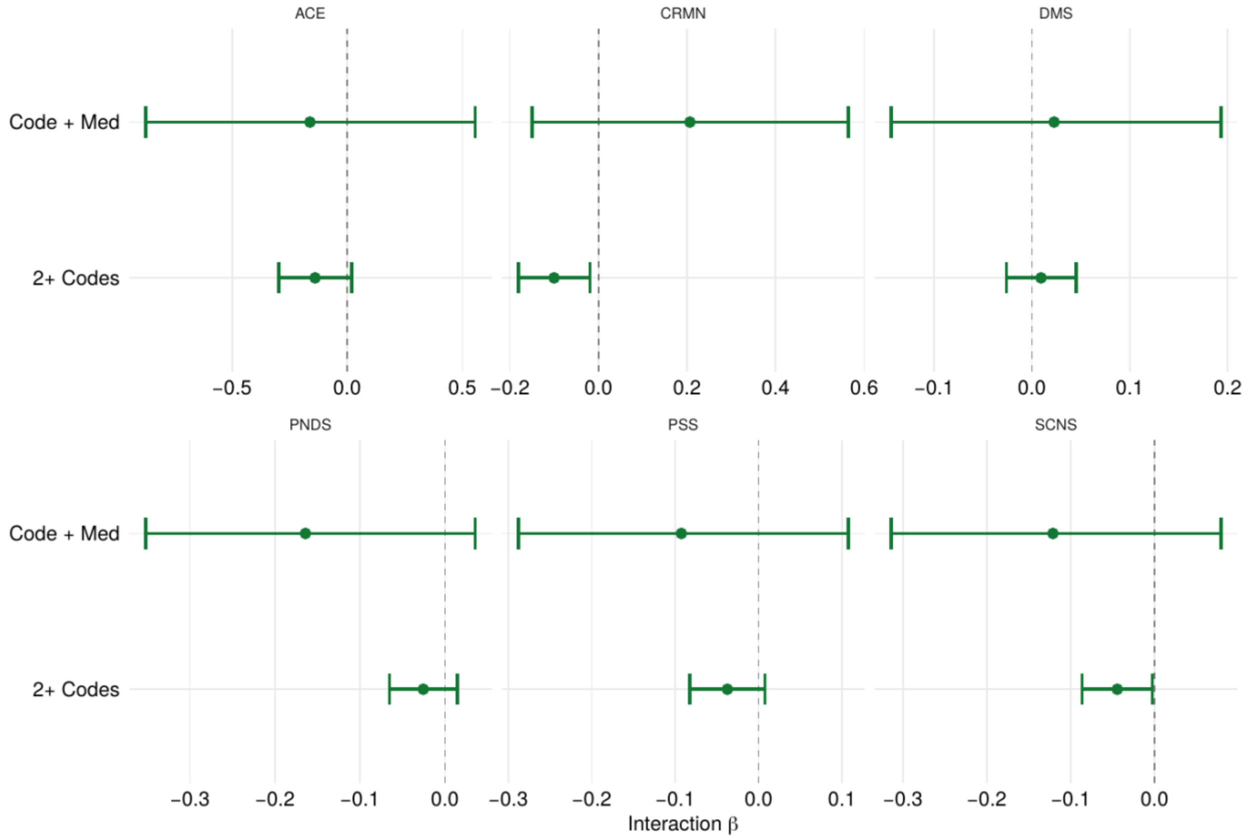
